# Associations of body fat and inflammation with non-communicable chronic diseases and mortality: A prospective cohort study

**DOI:** 10.1101/2025.06.03.25328903

**Authors:** Natasha Wiebe, Marcello Tonelli

**Author notes:** No reprints will be available. Correspondence to: Natasha Wiebe, University of Alberta 11-112Y CSB, 11350 83 Ave NW, Edmonton AB, T6G 2G3 Canada. **Conflict of Interest Disclosures:** None reported. **Funding:** David Freeze Chair in Health Services Research at the University of Calgary.

## Abstract

**Introduction:** Certain leading medical organizations are considering alternatives to the body mass index (BMI) as a predictor of the risk for non-communicable chronic disease (NCD) or death. Our objective was to evaluate the associations between various measures of body fat and the risk of incident NCDs or mortality, independent of inflammation.

**Methods:** This was a population-based prospective cohort study (the UK Biobank cohort) set in the United Kingdom. The participants (between 40 and 69 years) accrued between March 2006 and October 2010, and followed until December 2022. The exposures were BMI, waist:hip ratio (WHR), bioimpedance analysis-measured body fat (fat_BIA_), C-reactive protein (CRP) and various other measures of body fat obtained by dual-energy X-ray absorptiometry (including visceral adipose tissue) and magnetic resonance imaging. The outcomes were all-cause death, cardiovascular disease (heart failure, hypertension, myocardial infarction, pulmonary embolism, and stroke), cancers (breast, colorectal, endometrial, esophageal, kidney, ovarian, pancreatic, and prostate), diabetes, asthma, gallbladder disease, chronic back pain, and osteoarthritis.

**Results:** There were 500,107 participants: the median age was 58 years (interquartile range 50-63) at baseline and 45.6% were male. The 5^th^ and 95^th^%iles for measures of body fat were: BMI 20.5 (considered “healthy”) and 37.0 kg/m^2^ (considered “unhealthy”), WHR 0.71 and 0.94, BIA 24.8 and 47.6% in females, and in males BMI 22.0 (considered “healthy”) and 35.4 kg/m^2^ (considered “unhealthy”), WHR 0.83 and 1.05, BIA 15.5 and 34.7%. BMI was strongly correlated to fat_BIA_ (0.85 in females, 0.80 in males) but less so with WHR (0.46 in females, 0.59 in males).

All measures of body fat were positively associated with the incidence of NCDs but only WHR remained positively associated with death after full adjustment (HR 95^th^%ile vs 5^th^%ile [95%CI]: BMI 0.80 [0.76,0.84], WHR 1.21 [1.16,1.28], BIA 0.80 [0.76,0.84] in females; BMI 0.89 [0.85,0.93], WHR 1.19 [1.14,1.24], BIA 0.89 [0.85,0.92] in males). Simpler models that adjusted for age, sex, CRP, WHR and either BMI or fat_BIA_ gave similar results. Associations between body fat and the incidence of NCDs after accounting for the competing risk of death were also similar.

**Conclusion:** BMI was strongly correlated with fat_BIA_, but WHR and VAT_DXA_ were less so. All measures of body fat were associated with the incidence of NCDs but only WHR was independently associated with mortality. These findings support the hypothesis that body fat may be protective against death, and that the excess risk associated with higher WHR may be mediated by something other than body fat.

**Key Questions:** *What is already known about this topic?:*

- Obesity as measured by body mass index is associated with chronic disease but less so with mortality.
- Less is known about alternative measures of body fat.

*What this study adds?:*

- Body mass index is strongly correlated with body fat as measured by bioimpedance analysis, but less so with the waist:hip ratio and visceral adipose tissue.
- All measures of body fat were associated with the incidence of chronic disease but only the waist:hip ratio was independently associated with mortality.

*How this study might affect research, practice or policy?:*

- These findings support the hypothesis that body fat may be protective against death, and that the excess risk associated with higher waist:hip ratio may be mediated by something other than body fat.

## Introduction

Obesity is associated with increased prevalence of non-communicable chronic disease (NCD),(1) particularly type 2 diabetes, cardiovascular and pulmonary disease, and cancer.(2, 3) However, obesity (as assessed by high body mass index [BMI]) is not associated with excess mortality in many clinical populations.(4) In fact, after adjustment for key confounders such as inflammation and/or fasting insulin, higher BMI was associated with a lower risk of mortality in a representative population of US adults.(5)

Although both inflammation and hyperinsulinemia are associated with excess risk of incident NCDs, neither has been shown to be caused by obesity.(6) However, there is some evidence from animal studies that chronic elevations of C-reactive protein (CRP) may precede or even cause adult-onset obesity and possibly insulin resistance (IR).(7) Since adiposity plays a role in our immune system (i.e., apoptotic cell clearance, extracellular matrix remodeling, angiogenesis),(8) obesity may arise as part of a protective response to disease development.

Most studies examining the association between obesity and adverse clinical outcomes have relied on the BMI as a marker of body fat. However, several organizations such as the American Medical Association(9) are recommending consideration of alternatives to BMI including visceral fat, body adiposity index, body composition, relative fat mass, and waist circumference.

While many systematic reviews have noted that mortality is lower for people with higher BMI (the “obesity paradox”),(4, 10, 11, 12, 13, 14, 15, 16, 17, 18, 19, 20, 21, 22, 23, 24) few if any studies evaluate whether body fat (however measured), may reduce the risk of incident NCDs after accounting for inflammation. In this prospective cohort study of UK adults, we evaluated the associations of various body fat measures, independent of inflammation, with the development of NCDs and all-cause mortality. Given the potential importance of survivorship bias for analyses of non-fatal outcomes such as incident NCDs, we repeated all analyses using a competing risks framework.

## Methods

We report this prospective cohort study according to the STROBE guidelines.(25) All participants have signed written informed consent forms (National Research Ethics Service, reference 11/NW/0382). The institutional review boards at the Universities of Alberta (Pro00134819) and Calgary (pSite-23-0044) approved this study. As this study was a secondary analysis, it was not possible to include patients in the design and conduct of this study.

### Data sources and cohort

We used the UK Biobank database (www.ukbiobank.ac.uk), which incorporates data from over 500,000 adults between the ages of 40 and 69 years in England, Scotland and Wales. The UK Biobank includes data on demographics, social variables and behaviours, baseline medical conditions, imaging, prescriptions and supplements, biological samples, surgeries, genomics, and clinical outcomes. We used the database to assemble a cohort of adults with at least one measure of body fat. Participants were accrued between March 13, 2006 and October 1, 2010, and were followed until death, loss to follow-up or study end (December 31, 2022), whichever was earliest.

### Body fat and markers of inflammation

We obtained measures of baseline body fat via dual-energy X-ray absorptiometry (DXA; total tissue fat percentage [fat_DXA_] and visceral adipose tissue [VAT_DXA_] percentage), abdominal magnetic resonance imaging (MRI; subcutaneous adipose tissue [SAT_MRI_] percentage, VAT_MRI_ percentage, total abdominal adipose tissue index [index_MRI_; defined as (VAT_MRI_ volume + SAT_MRI_ volume)/height squared]), bioimpedance analysis (BIA; body fat percentage [fat_BIA_]), and physical measurement (BMI, waist:hip ratio [WHR]). Available markers of inflammation were: CRP, neutrophil-to-lymphocyte ratio (NLR), and platelet-to-lymphocyte ratio (PLR). Red cell distribution width (RDW), apolipoprotein B and glycated hemoglobin, all of which are also related to inflammation,(26, 27, 28) were obtained at baseline.

### Outcomes

In addition to all-cause mortality, we studied obesity-related NCDs identified by a systematic review by Guh *et al*(2) from 2009. Their review found evidence linking obesity with 18 NCDs: five cardiovascular diseases (specifically heart failure, hypertension, myocardial infarction, pulmonary embolism, stroke), eight cancers (specifically breast, colorectal, endometrial, esophageal, kidney, ovarian, pancreatic, prostate), type II diabetes (expanded in the current study to include type 1 diabetes), asthma, gallbladder disease, chronic back pain, and osteoarthritis. Death and cancer incidence were defined using available registries. Asthma, myocardial infarction and stroke were defined using validated administrative algorithms. The remaining seven outcomes were defined using the first incidence of a three-character International Classification of Diseases code obtained from primary care data, hospitalizations, the death registry, or self-reported conditions.

### Covariates

We obtained the following baseline variables: demographics (age, sex, ethnicity), social variables (rural/urban residence, Townsend deprivation index,(29) household income, employment status, education, social supports – friend and family visits, leisure/social activities, and the ability to confide in others), behaviours (smoking status, alcohol intake, physical activity [i.e., the International Physical Activity Question; IPAQ(30)], fruit and vegetable intake, sleep duration), comorbidities and other health related measures (walking pace, forced expiratory volume in 1 second, hand grip strength [maximum of left and right hands], and some form of a government accommodated disability). Comorbidities were Guh’s obesity-related conditions(2) (cardiovascular disease, cancer, diabetes, asthma, osteoporosis, gallbladder disease, and chronic back pain), and self-reported insomnia. The Townsend deprivation index incorporates unemployment, lack of car and home ownership, and household overcrowding.

Higher scores indicate more deprivation. Gender was not available.

### Statistical analyses

We did analyses with Stata MP 16·0 (www.stata.com) and report baseline descriptive statistics as counts and percentages, or medians and interquartile ranges, as appropriate. We tested differences across BMI groups using Chi-square and Kruskal-Wallis tests. We set the threshold *p* for statistical significance at 0.05. We calculated all pairwise correlations between the various body fat measures and with CRP by sex.

We used Cox regression to determine the associations between measures of body fat and time to clinical outcomes. In initial exploratory regressions with mortality, we determined that variables were best parametrized as follows: 1) quadratic and linear terms for body fat and most labs, and 2) a natural logarithmic term for CRP (lnCRP). Since quantities of body fat are expected to differ between the sexes, we interacted sex with each body fat measure. We report age-sex-lnCRP adjusted associations (primary analysis), and fully adjusted associations (using all the covariates mentioned above except for IPAQ and walking pace, which may be influenced by a participant’s weight). We determined that the proportional hazard assumption was satisfied by examining plots of the log-negative-log of within-group survivorship probabilities versus log-time after adjustment for covariates. Missingness of each variable is reported in Supplement Table S1. No outcomes or measures of body fat were imputed but missing values for covariates were imputed in the regressions: median value for continuous variables and most frequent value for categorical variables.

We report percentile-based hazard ratios (the 95^th^%ile vs 5^th^%ile) for the measures of body fat with corresponding 95% confidence intervals (CIs). The 5^th^%ile of BMI fell within the normal range of BMI for both females and males (i.e., 20.5 and 22.0 kg/m^2^) and the 95^th^%ile of BMI fell within the moderate obesity range for females and males (i.e., 37.0 and 35.5 kg/m^2^).

Additionally, we plotted continuous hazard ratios across the 1^st^ to the 99^th^ percentile range (referent at the 5^th^%ile) in the natural units of four body fat measures (BMI, WHR, fat_BIA_, and VAT_DXA_) superimposed on histograms showing the population distribution of each measure of body fat.

In sensitivity analyses, 1) we explored the independent effects of various subsets of covariates (i.e., age-sex, social determinants, behaviours, labs, and comorbidities and other health related measures) on the risk of outcomes, 2) we modelled mortality as a competing risk using a flexible modelling approach outlined by Lambert(31) (Weibull hazards regression with three or four knots), and 3) we adjusted for WHR together with BMI or fat_BIA_ to explore the independent associations of WHR from BMI and fat_BIA_.

## Results

Of 502,178 participants, 500,107 had a measure of body fat and thus are included in this study. Median follow-up time was 13.8 years (range 4 days to 16.8 years). There were 44,145 (8.8%) deaths, 76,982 (21.6%) new diagnoses of CVD, 33,088 (6.8%) first diagnoses of cancer, 25,384 (5.4%) new diagnoses of diabetes, 13,182 (3.0%) new diagnoses of asthma, 21,070 (4.4%) new diagnoses of gallbladder disease, 35,500 (7.8%) new diagnoses of chronic back pain, and 40,133 (9.0%) new diagnoses of osteoarthritis.

Supplement Table S1 summarizes demographics and clinical characteristics by BMI categories: <18.5, 18.5-24.9, 25.0-34.9, and ≥35.0 kg/m^2^. The participants in the highest weight category were the most deprived according to the Townsend index, had the lowest income, did the most unpaid voluntary work, were most likely to be non-white, and to have some kind of disability accommodation. They were most likely to have quit smoking and to drink only on special occasions, had the most friend or family visits but felt the least likely to be able to confide. They had the slowest walking pace and the lowest IPAQ scores (although neither variable accounted for a participant’s weight). They had the most insomnia and the most of every obesity-related comorbidity. They had the highest CRP, RDW and glycated haemoglobin, and they had the highest measures of body fat on all measures except for VAT_MRI_.

The participants in the lowest weight category were the most likely to be unemployed, a homemaker or on disability income. They had the fastest walking pace and were most likely to not drink alcohol but were the most likely to smoke. They had the fewest visits from friends and family, and they were the least likely to participate in social activities. They had the highest NLRs and PLRs.

There were a number of strong correlations between body fat measures (Table 1). Fat_BIA_ and index_MRI_ were both strongly correlated with BMI (0.85 in females and 0.80 in males, and 0.82 and 0.79, respectively). Fat_DXA_ was very strongly correlated with the index_MRI_ (0.91 in females, and 0.92 in males). Lastly, VAT_MRI_ was strongly correlated with VAT_DXA_ (0.84 in females, and 0.78 in males). SAT_MRI_ and VAT_MRI_ were dropped from further analyses due to lower power (sample sizes <5% of the total).

**Table 1.** Correlations of body fat and C-reactive protein by sex.

|  | BMI | WHR | Fat<sub>BIA</sub> | Fat<sub>DXA</sub> | VAT<sub>DXA</sub> | Index<sub>MRI</sub> | SAT<sub>MRI</sub> | VAT<sub>MRI</sub> | lnCRP |
| --- | --- | --- | --- | --- | --- | --- | --- | --- | --- |
| BMI | - | 0.59 | **0.80** | 0.66 | 0.59 | 0.79 | 0.35 | 0.32 | 0.33 |
| WHR | 0.46 | - | 0.62 | 0.55 | 0.60 | 0.60 | 0.17 | 0.46 | 0.37 |
| Fat<sub>BIA</sub> | **0.85** | 0.46 | - | 0.74 | 0.62 | 0.75 | 0.31 | 0.34 | 0.38 |
| Fat<sub>DXA</sub> | 0.70 | 0.35 | 0.78 | - | **0.80** | **0.92** | 0.35 | 0.46 | 0.37 |
| VAT<sub>DXA</sub> | 0.58 | 0.57 | 0.59 | 0.71 | - | **0.85** | 0.03 | 0.78 | 0.35 |
| Index<sub>MRI</sub> | **0.82** | 0.47 | 0.78 | **0.91** | 0.79 | - | 0.37 | 0.53 | 0.37 |
| SAT<sub>MRI</sub> | 0.48 | 0.39 | 0.49 | 0.57 | 0.41 | 0.66 | - | -0.28 | 0.09 |
| VAT<sub>MRI</sub> | 0.32 | 0.58 | 0.37 | 0.46 | **0.84** | 0.54 | 0.26 | - | 0.23 |
| lnCRP | 0.50 | 0.34 | 0.52 | 0.45 | 0.41 | 0.48 | 0.30 | 0.34 | - |

BIA bioimpedance analysis, BMI body mass index, DXA dual-energy X-ray absorptiometry, lnCRP natural logarithm of C-reactive protein, MRI magnetic resonance imaging, SAT subcutaneous adipose tissue, VAT visceral adipose tissue, WHR waist:hip ratio
Data from females are shown in the lower triangle and data from males are shown in the upper triangle. Correlations are bolded if $\geq 0.80$ .

In females, the correlations between body fat measures and lnCRP tended to be around 0.41 to 0.50, except for WHR which was 0.34. In males, these correlations tended to be around 0.33-0.38, whereas for WHR it was 0.37.

Supplement Table S2 shows all the age-sex-lnCRP adjusted associations between body fat and outcomes. Figure 1 is an abbreviated version of Supplement Table S2 (together with results of the competing risk analyses). Despite adjustment for lnCRP, almost all measures of body fat were positively associated with every obesity-related NCD (where power allowed) in both sexes (Supplement Figures S1-S7). Diabetes (HRs from 4.43 to 16.32) then gallbladder disease (HRs from 2.75 to 7.16) were the most strongly associated with a measure of body fat.

**Figure 1.**
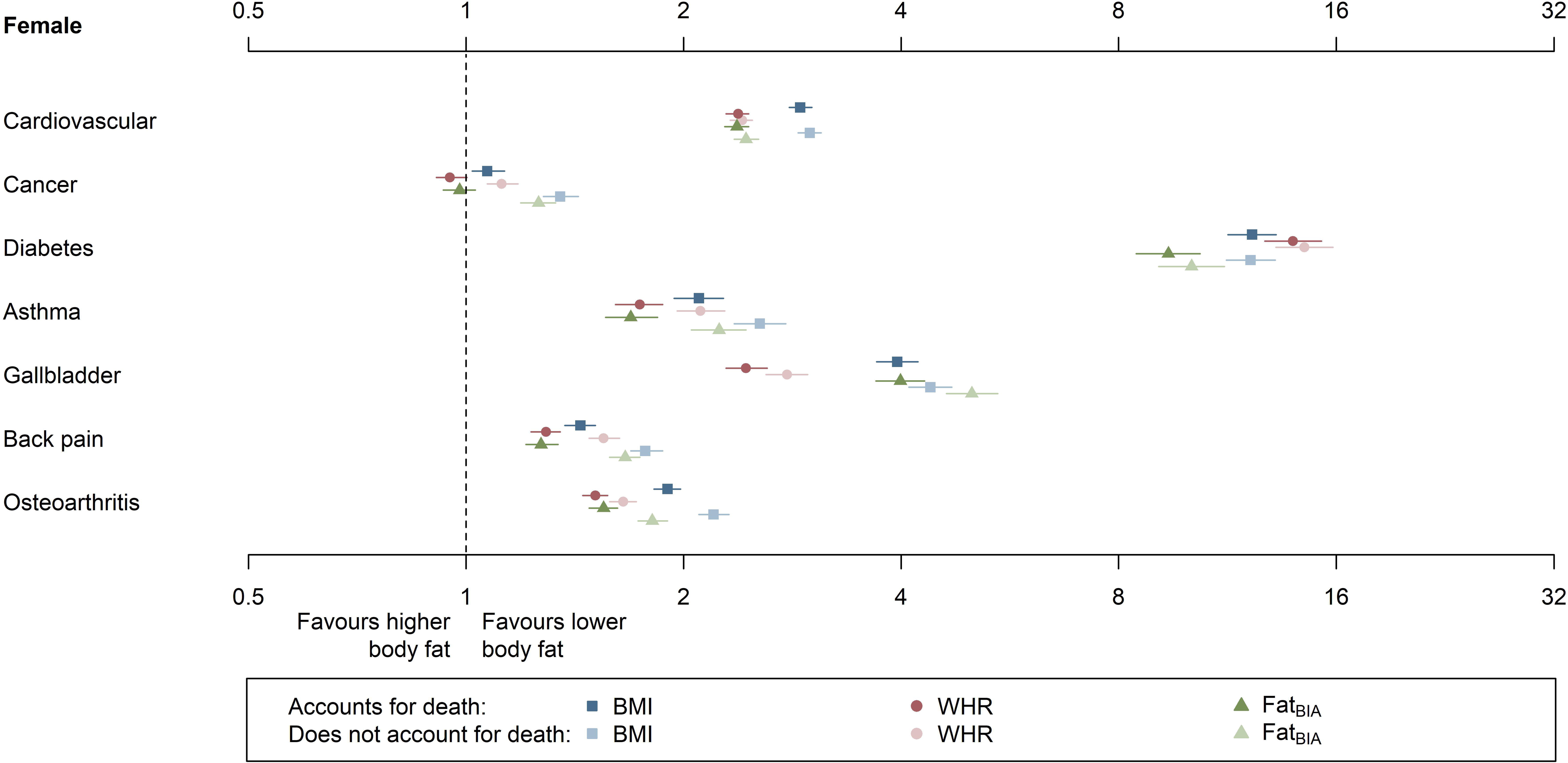
Associations of body fat measures with clinical outcomes with and without the competing risk of death. BIA bioimpedance analysis, BMI body mass index, HR hazard ratio, lnCRP natural logarithm of C-reactive protein, SHR subdistribution hazard ratio, WHR waist:hip ratio Results from six models are presented above (3 measures of body fat X accounting or not accounting for death). Each marker is a HR or SHR comparing the 95^th^%ile versus the 5^th^%ile of a body fat measure. The coloured horizontal lines represent the 95% confidence limits. The models are adjusted for age, sex, lnCRP and one of three measure of body fat (linear and quadratic terms). Sex is interacted with each measures of body fat. The faded markers do not adjust for the competing risk of death. The non-faded markers account for the competing risk of death.

The histograms in Supplement Figures S1-S7 (and Figure 2) show that females tended to have smaller WHRs and smaller quantities of VAT_DXA_ but larger quantities of fat_BIA_ than males. The distribution of BMI was quite similar between the sexes although the distribution for females was wider. In contrast to the reporting of 95^th^ versus 5^th^ percentile HRs (where comparisons using percentiles offset any between-sex differences in absolute body fat), these figures using the natural units show that females were at higher risk for NCDs than males at lower WHRs and VAT_DXA_ but were at lower risk with higher fat_BIA_. Risk was similar between females and males for comparable levels of BMI.

**Figure 2.**
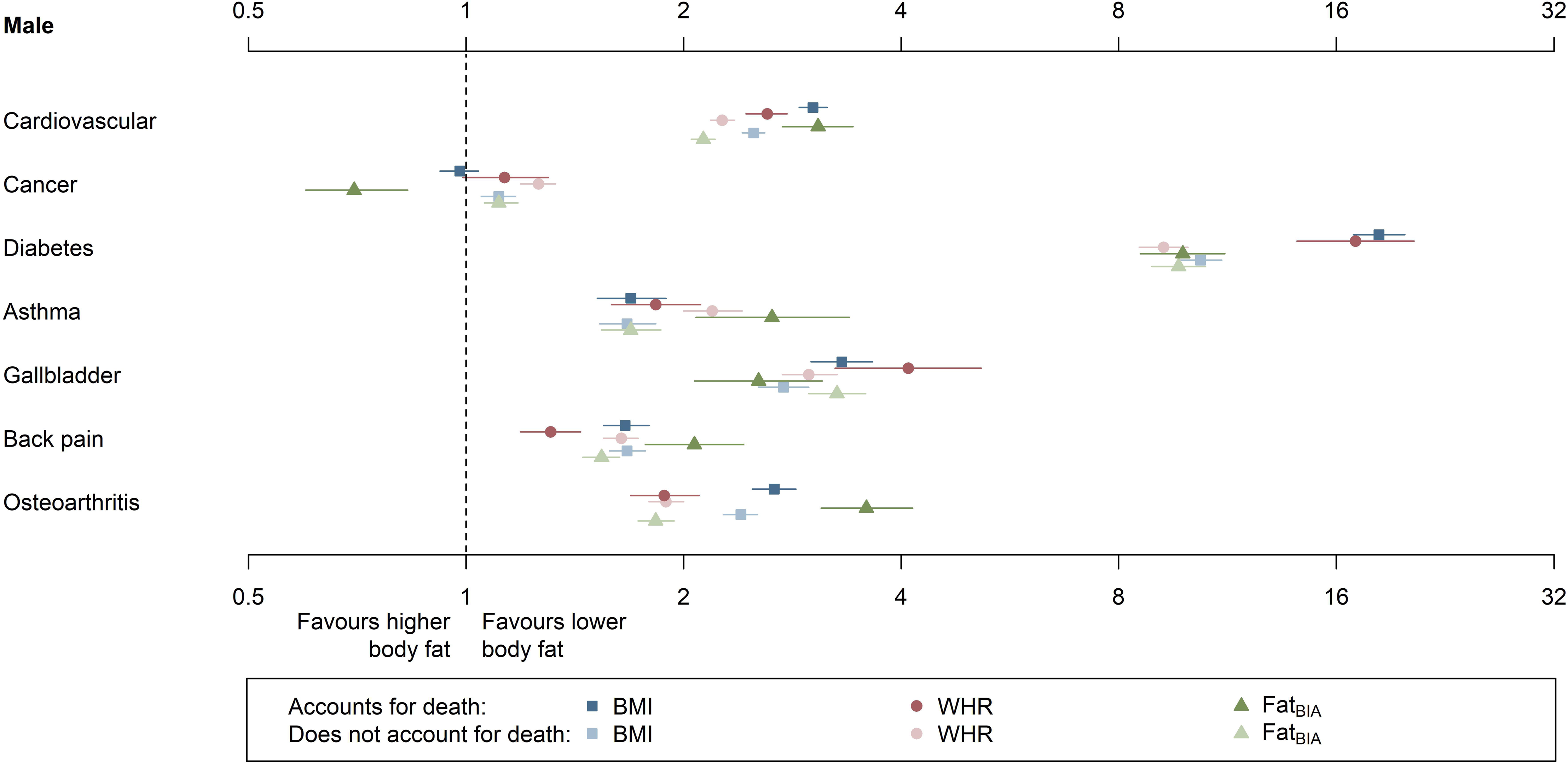
Associations of body fat measures with all-cause mortality BIA bioimpedance analysis, BMI body mass index, DXA dual-energy X-ray absorptiometry, HR hazard ratio, lnCRP natural logarithm of C-reactive protein, VAT visceral adipose tissue, WHR waist:hip ratio Results from four models are presented above (4 measures of body fat). Plots show hazard ratios plotted against BMI (top-left), WHR (top-right), fat_BIA_ (bottom-left), and VAT_DXA_ (bottom-right). Curves in pink represent females and curves in blue represent males. Solid lines represent HR adjusted for age, sex, and lnCRP; and dashed lines represent fully adjusted HR. The range of body fat represent the 3^rd^%ile to the 97^th^%ile within sex. Histograms underlaying the HR plots show the distribution of the various measures of body fat by sex (pink for females, blue for males, and purple for overlapping distributions).

The age-sex-lnCRP adjusted findings for mortality were not consistent across measures of body fat, nor between sexes (Figure 2, Supplement Table S2). The DXA and MRI measures of body fat were not significantly associated with mortality in either sex, perhaps because of inadequate statistical power due to their much lower sample size (∼10% of the total N). These measures were dropped from further analyses. In females, WHR was positively associated (HR 1.69, 95% CI 1.61,1.77), BMI was not significantly associated (HR 0.98, 95% CI 0.93, 1.03), and fat_BIA_ was negatively associated with mortality (HR 0.93, 95% CI 0.89, 0.98). In males, all three – WHR, BMI and fat_BIA_ – were positively associated with mortality (HR 1.91, 95% CI 1.83,1.99; HR 1.11, 95% CI 1.07,1.16; HR 1.17, 95% CI 1.12,1.22).

After full adjustment (Figure 3, Supplement Table S3), all measures of body fat remained positively associated with all obesity-related NCDs. In contrast, BMI and fat_BIA_ were negatively associated with mortality in both sexes (HR 0.80, 95% CI 0.76,0.84 and HR 0.80, 95% 0.76,0.84 in females; HR 0.89, 95% CI 0.85,0.93 and HR 0.89, 95% CI 0.85,0.92 in males). However, WHR remained positively associated with mortality in both sexes (HR 1.21, 95% CI 1.16,1.28 in females; HR 1.19, 95% CI 1.14,1.24 in males). In sensitivity analyses with adjustment for various subsets of covariates (Figure 3, Supplement Table S3), the laboratory measures and the comorbidities/health related measures appeared to have the largest attenuating effects on the associations with mortality.

**Figure 3.**
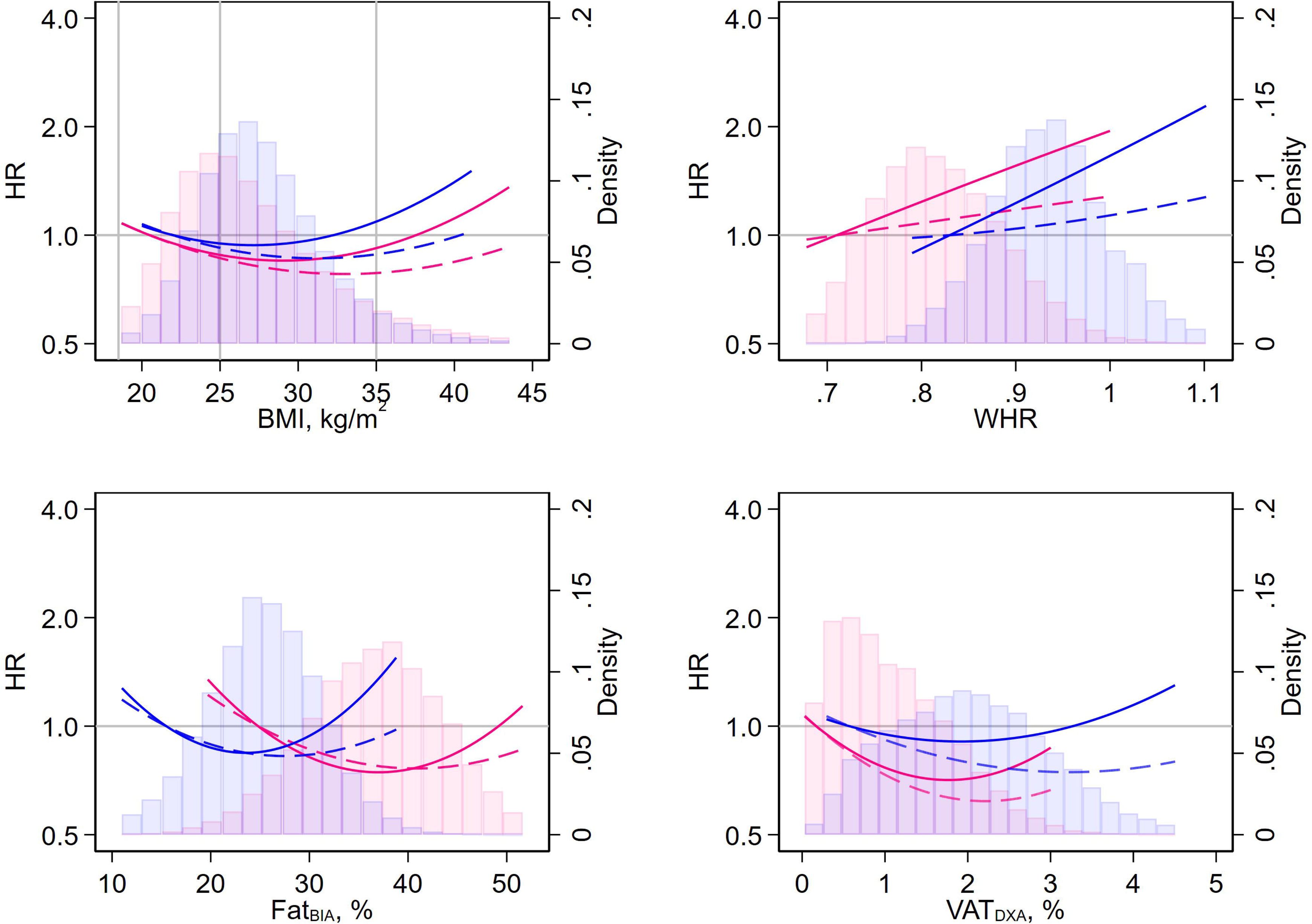
Associations of body fat measures with clinical outcomes adjusted for various sets of covariates BIA bioimpedance analysis, BMI body mass index, HR hazard ratio, IPAQ International Physical Activity Questionnaire, lnCRP natural logarithm of C-reactive protein, NLR neutrophil:lymphocyte ratio, PLR platelet:lymphocyte ratio, RDW red cell distribution width, WHR waist:hip ratio Results from 18 models are presented above (3 measures of body fat X 6 sets of covariates). Each marker is a HR comparing the 95^th^%ile versus the 5^th^%ile of a body fat measure. The coloured horizontal lines represent the 95% confidence limits. Each model is adjusted for one of the three measures of body fat: BMI, WHR, or fat_BIA_. The model represented with the black asterisks adjusted for all covariates listed in Table 1 except for IPAQ and walking pace which may be determined, at least in part, by a participant’s body fat. The model represented with the blue squares adjusted for age and sex but not lnCRP. The model represented with the yellow circles adjusted for age, sex, and social variables (Townsend deprivation index, rural residence, household income, employment, post-secondary education, non-white ethnicity, friends and family visits, leisure/social activities, able to confide). The model represented with the orange triangles adjusted for age, sex, and behaviours (alcohol consumption, fresh fruit consumption, sleep duration, smoking status, raw vegetable consumption). The model with the maroon diamonds adjusted for age, sex, and labs (lnCRP, apolipoprotein B, glycated hemoglobin, NLR, PLR, RDW - all quadratic terms except CRP). The model represented with the upside down green triangle adjusted for age, sex, and baseline comorbidities (cardiovascular disease, cancer, diabetes, asthma, osteoarthritis, gallbladder disease, chronic back pain) and other related health measures (forced expiratory volume in 1s, hand grip strength, government-accommodated disability, insomnia).

When accounting for the competing risk of death (Figure 1, Supplement Table S4), most of the associations between body fat and the risk of incident NCDs were attenuated slightly in females, but were somewhat amplified in males, especially for diabetes. In contrast, accounting for the competing risk of death largely abrogated the associations between measures of body fat and incident cancer for both sexes.

After adjustment for WHR and BMI or fat_BIA_, the associations between body fat and mortality remained different from those between body fat and the risk of incident NCDs (Figure 4, Supplement Table S5). WHR maintained positive associations with mortality but in these analyses BMI (females: HR 0.75, 95% CI 0.71,0.79; males: HR 0.67, 95% CI 0.64,0.71) and fat_BIA_ (females: HR 0.73, 95% CI 0.69,0.77; males: HR 0.78, 95% CI 0.74,0.82) were negatively associated with mortality in both females and males.

**Figure 4.**
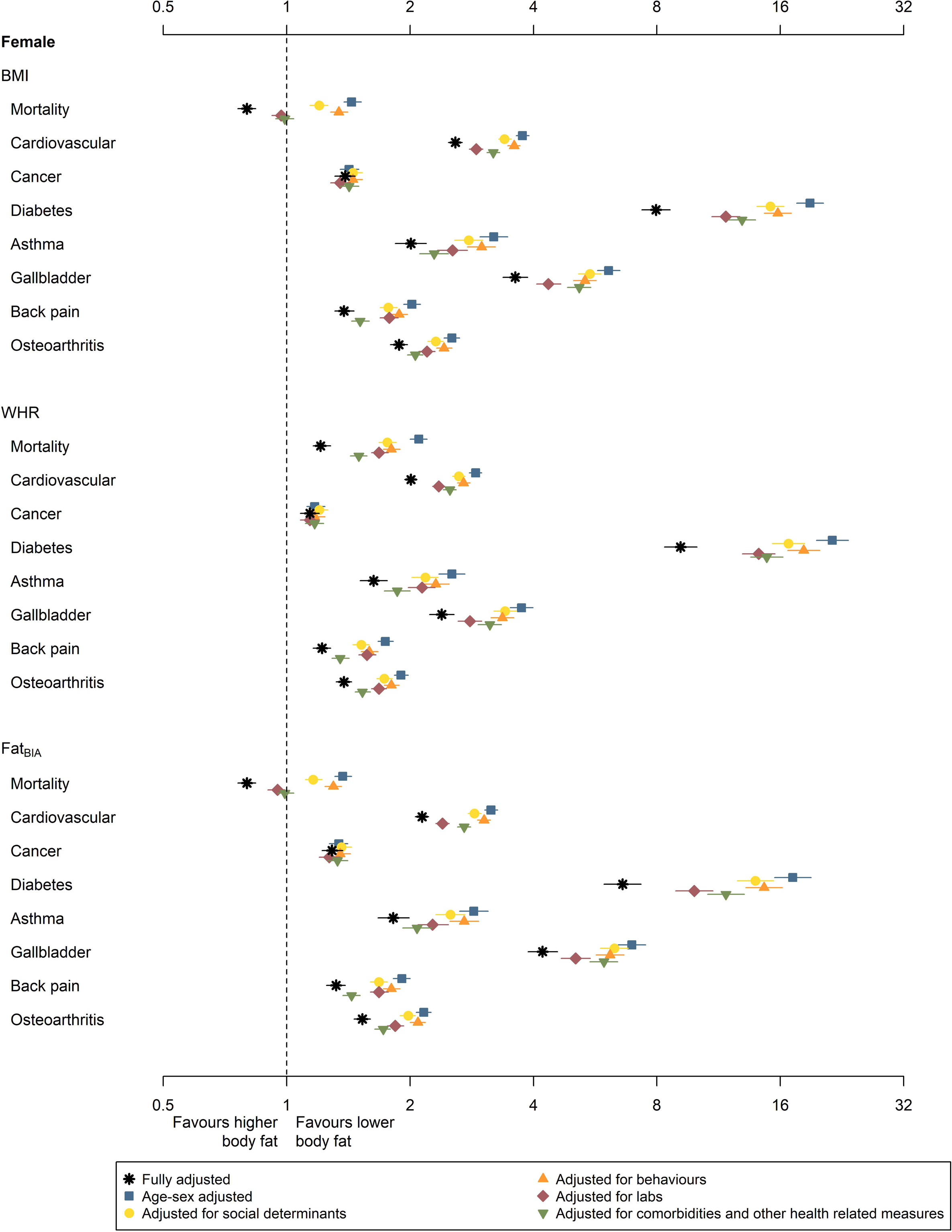
Associations of body fat measures with clinical outcomes adjusted for two measures of body fat BIA bioimpedance analysis, BMI body mass index, HR hazard ratio, lnCRP natural logarithm of C-reactive protein, WHR waist:hip ratio Results from two model are presented above. Each marker is a HR comparing the 95^th^%ile versus the 5^th^%ile of a body fat measure. The coloured horizontal lines represent the 95% confidence limits. The models are adjusted for age, sex, lnCRP and two measures of body fat (linear and quadratic terms). The first model adjusts for BMI and WHR. The second model adjusts for fat_BIA_ and WHR. Sex is interacted with the measures of body fat.

## Discussion

We found that BMI was strongly correlated with body fat as assessed by BIA, but WHR and VAT_DXA_ were less so. All of these measures were positively associated with the incidence of NCDs despite adjustment for CRP and other available characteristics. In contrast, only WHR remained positively and significantly associated with all-cause mortality after adjustment for potential confounders. These findings support the hypothesis that body fat may be protective against death, and that the excess risk associated with higher WHR may be mediated by something other than body fat.

With or without adjustment for CRP, age, sex, social determinants, behaviours, other inflammatory-related labs, and baseline conditions and further health related measures, higher body fat was associated with a significantly increased risk of incident NCDs. The findings from competing risk analyses suggest that the latter is not due to survival bias, except perhaps for cancer where the associations between higher body fat and higher risk were no longer apparent. Similarly, our findings do not support a clinically meaningful effect of collider bias (where higher-weight patients may be screened and diagnosed more frequently than lower-weight patients), especially for males, where consideration of competing risks tended to amplify rather than attenuate the subdistribution hazard ratios for incident NCDs.

In contrast, the fully adjusted associations between measures of body fat and all-cause mortality were negatively associated when BMI or BIA was used to assess body fat, but was positively associated for WHR. Unlike WHR, the BIA-measured body fat as a predictor acted similarly to BMI throughout the analyses. Moreover, in age-sex-lnCRP adjusted models, WHR remained associated with higher mortality after adjustment for BMI or fat_BIA_, whereas BMI and fat_BIA_ were negatively associated with mortality after adjustment for WHR. There was no evidence that visceral fat as measured by DXA had any particular advantages for predicting the risk of mortality, although these analyses were underpowered (Figure 2).

Several studies(32, 33) have noted that WHR is more strongly associated with hyperinsulinemia and IR than is the BMI. Our previous work with the NHANES US study cohort(5) found that adjustment for fasting insulin was sufficient to uncover an association between higher BMI and lower mortality in a general population of US adults, particularly in males. Furthermore, four large studies (totalling ∼10,000 participants) in adults without diabetes(34, 35, 36, 37) show that rises in markers of inflammation precede increases in insulin level. Further, randomized trials(38, 39) and a meta-analysis(6) (with 60 studies) both show that changes in insulin levels precede changes in BMI. With this sequence in mind, it is possible that increased adiposity may blunt the hyperglycaemic stress response to NCD development and protect against death and potentially other consequences of NCDs.

Our study also has important limitations that should be considered when interpreting results. First, as with any observational study, residual confounding remains possible. We did not have data on weight cycling,(40, 41) weight stigma,(42, 43, 44) weight-based discrimination (e.g., less access to therapeutic surgeries and diagnostic imaging), and hyperinsulinemia/insulin resistance – all of which are more common among higher-weight patients. Second, the study population was not representative of the general UK population; 95% of participants were white and the proportion who died was higher in males than females (11.4% vs 6.7%).

Therefore our findings may be less generalizable to non-white people, and perhaps to healthier males. Third, while no study has established that body fat causes poor outcomes, some of the covariates in the fully adjusted analyses might mediate the associations between body fat and outcomes. Although we cannot rule out this possibility, mediation would be expected to bias towards the null (not past the null) and is unlikely to explain the finding that the association between mortality and higher BMI and fat_BIA_ inverts (shifts from positive to negative) following statistical adjustment, regardless of whether models include age, sex, lnCRP and WHR, or the full set of covariates.

In conclusion, BMI was strongly correlated with body fat as assessed by BIA, but WHR and VAT_DXA_ were less so. All measures of body fat were associated with the incidence of NCDs but only WHR was independently associated with mortality.

## Data Availability

These data are publicly available from https:// https://www.ukbiobank.ac.uk/.

## Acknowledgements

Anita Lloyd, MSc, PStat, University of Alberta performed a technical review; Sophanny Tiv, BSc, University of Alberta provided graphics support.

## Contributors

NW conceived the study, wrote the first draft of the manuscript, and performed the statistical analysis. NW and MT designed the study. Both authors critically reviewed, revised, and approved the final manuscript. NW had full access to all of the data in the study and takes responsibility for the integrity of the data and the accuracy of the data analysis.

## Disclaimer

This research has been conducted using the UK Biobank Resource under Application Number 144943. This work uses data provided by patients and collected by the NHS as part of their care and support. This study’s data is provided by the UK Biobank, NHS England and National Safe Haven. The interpretation and conclusions contained herein are those of the researchers and do not represent the views of the UK Biobank, NHS England or the National Safe Haven.

## Data Sharing Statement

These data are publicly available from https://https://www.ukbiobank.ac.uk/.

## Supplement legend

Supplement Figure S1. Associations of body fat measures with cardiovascular disease

Supplement Figure S2. Associations of body fat measures with cancer

Supplement Figure S3. Associations of body fat measures with diabetes

Supplement Figure S4. Associations of body fat measures with asthma

Supplement Figure S5. Associations of body fat measures with gallbladder disease

Supplement Figure S6. Associations of body fat measures with back pain

Supplement Figure S7. Associations of body fat measures with osteoarthritis

Supplement Table S1. Baseline demographics and clinical characteristics by body mass index

Supplement Table S2. Associations of body fat with mortality and other clinical outcomes by sex, HR (95% CI) – primary analysis – 95^th^%ile vs 5^th^%ile

Supplement Table S3. Associations of body fat with mortality and other clinical outcomes by sex, HR (95% CI) – covariate analyses – 95^th^%ile vs 5^th^%ile

Supplement Table S4. Associations of body fat with clinical outcomes, with mortality modelled as a competing risk by sex, SHR (95% CI) – 95^th^%ile vs 5^th^%ile

Supplement Table S5. Associations of BMI or fat_BIA_ and WHR with mortality and other clinical outcomes by sex, HR (95% CI) – 95^th^%ile vs 5^th^%ile

---

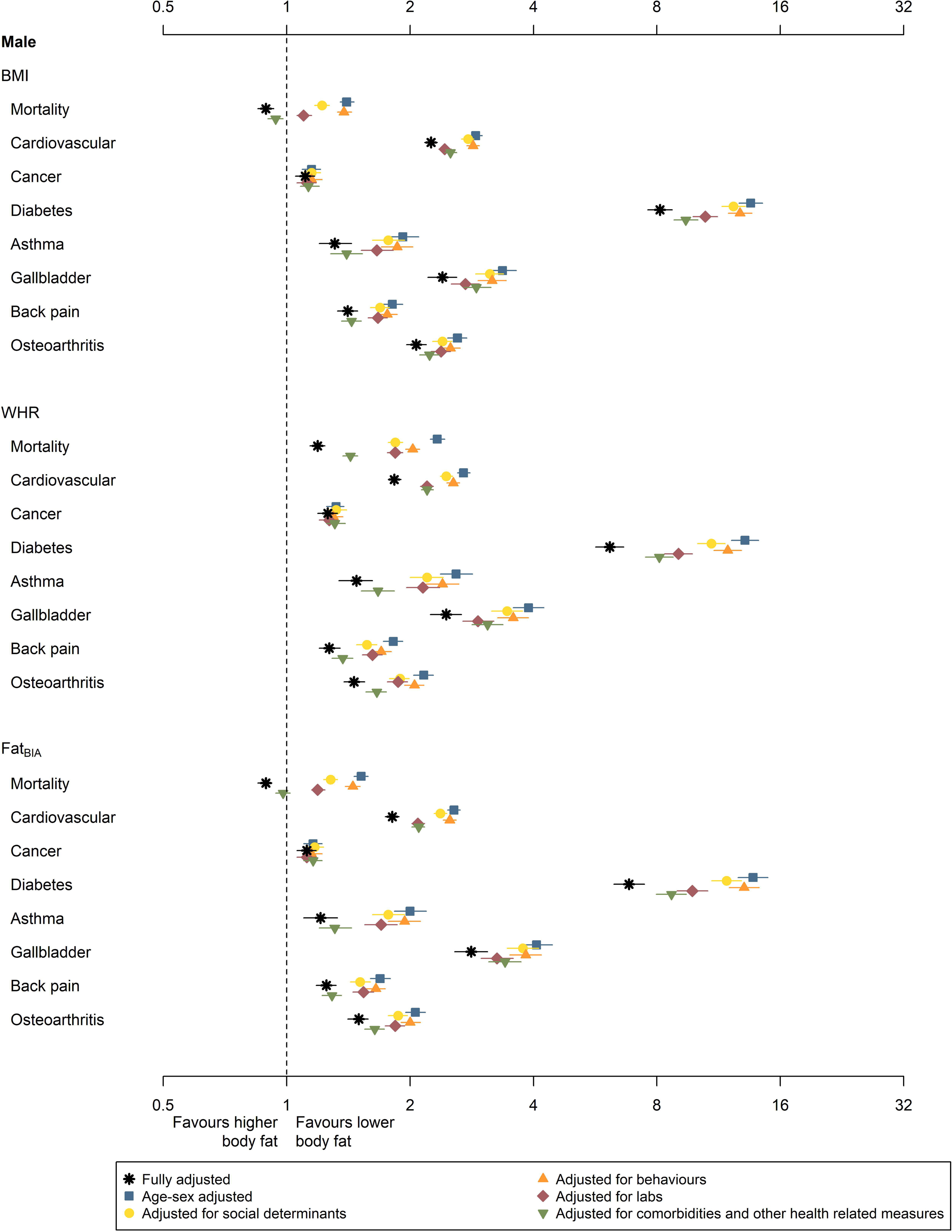

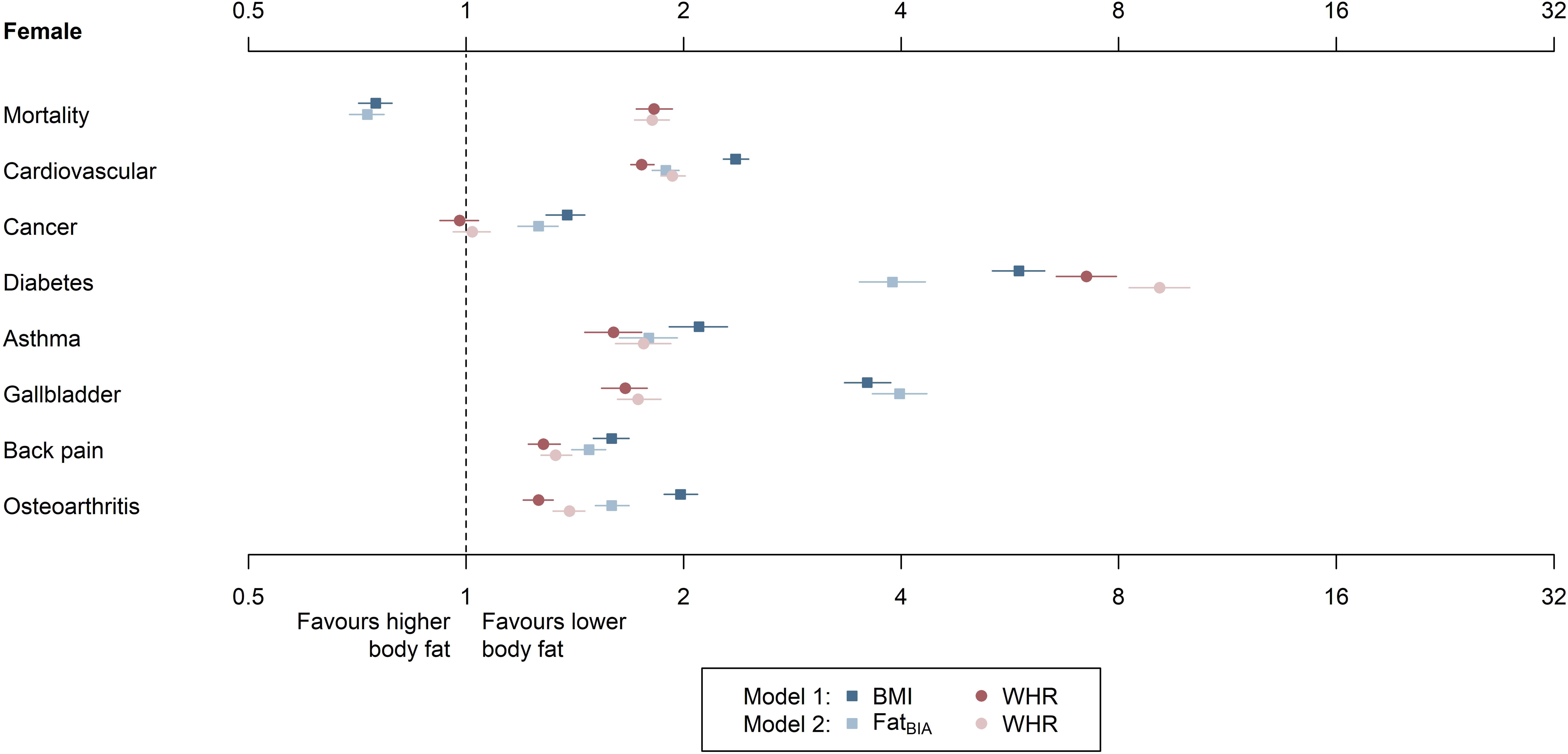

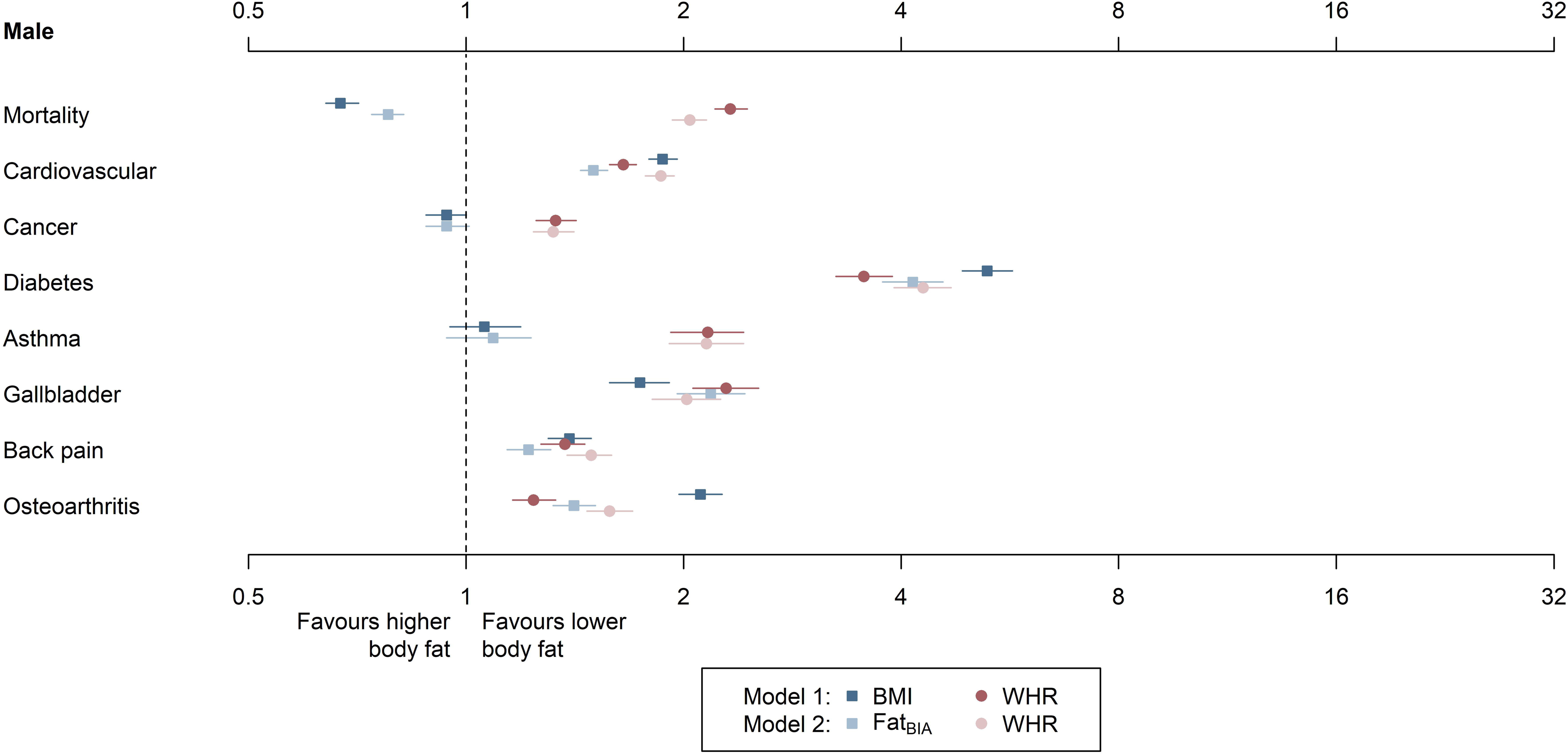

## References

1. Nyberg ST, Batty GD, Pentti J, Virtanen M, Alfredsson L, Fransson EI, et al. Obesity and loss of disease-free years owing to major non-communicable diseases: a multicohort study. Lancet Public Health 2018;3: e490–e497.

2. Guh DP, Zhang W, Bansback N, Amarsi Z, Birmingham CL, Anis AH. The incidence of co-morbidities related to obesity and overweight: a systematic review and meta-analysis. BMC Public Health 2009;9: 88.

3. Wiebe N, Stenvinkel P, Tonelli M. Associations of Chronic Inflammation, Insulin Resistance, and Severe Obesity With Mortality, Myocardial Infarction, Cancer, and Chronic Pulmonary Disease. JAMA Netw Open 2019;2: e1910456.

4. Wiebe N, Lloyd A, Crumley ET, Tonelli M. Associations between body mass index and all-cause mortality: A systematic review and meta-analysis. Obes Rev 2023: e13588.

5. Wiebe N, Muntner P, Tonelli M. Associations of body mass index, fasting insulin, and inflammation with mortality: a prospective cohort study. Int J Obes (Lond) 2022;46: 2107–2113.

6. Wiebe N, Ye F, Crumley ET, Bello A, Stenvinkel P, Tonelli M. Temporal Associations Among Body Mass Index, Fasting Insulin, and Systemic Inflammation: A Systematic Review and Meta-analysis. JAMA Netw Open 2021;4: e211263.

7. Li Q, Wang Q, Xu W, Ma Y, Wang Q, Eatman D, et al. C-Reactive Protein Causes Adult-Onset Obesity Through Chronic Inflammatory Mechanism. Front Cell Dev Biol 2020;8: 18.

8. Schipper HS, Prakken B, Kalkhoven E, Boes M. Adipose tissue-resident immune cells: key players in immunometabolism. Trends Endocrinol Metab 2012;23: 407–415.

9. American Medication Assocation. Support removeal of BMI as a standard measure in medicine and recognizing culturally-diverse and varied presentations of eating disorders and indications for metabolic and bariatric surgery: Report 09 of the Council on Science and Publish Health (A-23) (Reference Committee D): American Medication Assocation; 2020 [1 Sep 2023]. Available from: https://www.ama-assn.org/system/files/a23-csaph07.pdf.

10. Xie H, Wei L, Zhang H, Ruan G, Liu X, Lin S, et al. Association of systemic inflammation with the obesity paradox in cancer: results from multi-cohort studies. Inflamm Res 2024;73: 243–252.

11. Bundhun PK, Li N, Chen MH. Does an Obesity Paradox Really Exist After Cardiovascular Intervention?: A Systematic Review and Meta-Analysis of Randomized Controlled Trials and Observational Studies. Medicine (Baltimore) 2015;94: e1910.

12. Grymonprez M, Capiau A, De Backer TL, Steurbaut S, Boussery K, Lahousse L. The impact of underweight and obesity on outcomes in anticoagulated patients with atrial fibrillation: A systematic review and meta-analysis on the obesity paradox. Clin Cardiol 2021;44: 599–608.

13. Kwon Y, Kim HJ, Park S, Park YG, Cho KH. Body Mass Index-Related Mortality in Patients with Type 2 Diabetes and Heterogeneity in Obesity Paradox Studies: A Dose-Response Meta-Analysis. PLoS One 2017;12: e0168247.

14. Li S, Wang Z, Huang J, Fan J, Du H, Liu L, et al. Systematic review of prognostic roles of body mass index for patients undergoing lung cancer surgery: does the’obesity paradox’ really exist? Eur J Cardiothorac Surg 2017;51: 817–828.

15. Li Y, Li C, Wu G, Yang W, Wang X, Duan L, et al. The obesity paradox in patients with colorectal cancer: a systematic review and meta-analysis. Nutr Rev 2022;80: 1755–1768.

16. Lin GM, Li YH, Yin WH, Wu YW, Chu PH, Wu CC, et al. The Obesity-Mortality Paradox in Patients With Heart Failure in Taiwan and a Collaborative Meta-Analysis for East Asian Patients. Am J Cardiol 2016;118: 1011–1018.

17. Lv W, Li S, Liao Y, Zhao Z, Che G, Chen M, et al. The’obesity paradox’ does exist in patients undergoing transcatheter aortic valve implantation for aortic stenosis: a systematic review and meta-analysis. Interact Cardiovasc Thorac Surg 2017;25: 633–642.

18. Marcks N, Aimo A, Januzzi JL, Jr., Vergaro G, Clerico A, Latini R, et al. Re-appraisal of the obesity paradox in heart failure: a meta-analysis of individual data. Clin Res Cardiol 2021;110: 1280–1291.

19. Mei X, Hu S, Mi L, Zhou Y, Chen T. Body mass index and all-cause mortality in patients with percutaneous coronary intervention: A dose-response meta-analysis of obesity paradox. Obes Rev 2021;22: e13107.

20. Nie W, Zhang Y, Jee SH, Jung KJ, Li B, Xiu Q. Obesity survival paradox in pneumonia: a meta-analysis. BMC Med 2014;12: 61.

21. Niedziela J, Hudzik B, Niedziela N, Gasior M, Gierlotka M, Wasilewski J, et al. The obesity paradox in acute coronary syndrome: a meta-analysis. Eur J Epidemiol 2014;29: 801–812.

22. Padwal R, McAlister FA, McMurray JJ, Cowie MR, Rich M, Pocock S, et al. The obesity paradox in heart failure patients with preserved versus reduced ejection fraction: a meta-analysis of individual patient data. Int J Obes (Lond) 2014;38: 1110–1114.

23. Tan XF, Shi JX, Chen AM. Prolonged and intensive medication use are associated with the obesity paradox after percutaneous coronary intervention: a systematic review and meta-analysis of 12 studies. BMC Cardiovasc Disord 2016;16: 125.

24. Zhi G, Xin W, Ying W, Guohong X, Shuying L. “Obesity Paradox” in Acute Respiratory Distress Syndrome: Asystematic Review and Meta-Analysis. PLoS One 2016;11: e0163677.

25. von Elm E, Altman DG, Egger M, Pocock SJ, Gotzsche PC, Vandenbroucke JP, et al. The Strengthening the Reporting of Observational Studies in Epidemiology (STROBE) statement: guidelines for reporting observational studies. Lancet 2007;370: 1453–1457.

26. Lippi G, Targher G, Montagnana M, Salvagno GL, Zoppini G, Guidi GC. Relation between red blood cell distribution width and inflammatory biomarkers in a large cohort of unselected outpatients. Arch Pathol Lab Med 2009;133: 628–632.

27. Liu S, Hempe JM, McCarter RJ, Li S, Fonseca VA. Association between Inflammation and Biological Variation in Hemoglobin A1c in U.S. Nondiabetic Adults. J Clin Endocrinol Metab 2015;100: 2364–2371.

28. Faraj M, Messier L, Bastard JP, Tardif A, Godbout A, Prud’homme D, et al. Apolipoprotein B: a predictor of inflammatory status in postmenopausal overweight and obese women. Diabetologia 2006;49: 1637–1646.

29. Townsend P. Deprivation. J Soc Pol 1987;16: 125–146.

30. IPAQ. Guidelines for data processing and analysis of the International Physical Activity Questionniare (IPAQ) - short and long forms 2005 [30 May 2024]. Available from: https://biobank.ndph.ox.ac.uk/showcase/ukb/docs/ipaq_analysis.pdf.

31. Lambert PC. The estimation and modelling of cause-specific cumulative incidence functions using time-dependent weights. Stata J 2017;17: 181–207.

32. Benites-Zapata VA, Toro-Huamanchumo CJ, Urrunaga-Pastor D, Guarnizo-Poma M, Lazaro-Alcantara H, Paico-Palacios S, et al. High waist-to-hip ratio levels are associated with insulin resistance markers in normal-weight women. Diabetes Metab Syndr 2019;13: 636–642.

33. Liu MM, Liu QJ, Wen J, Wang M, Liang-Yan W, Qu ML, et al. Waist-to-hip ratio is the most relevant obesity index at each phase of insulin secretion among obese patients. J Diabetes Complications 2018;32: 670–676.

34. Yan YK, Li SX, Liu Y, Bazzano L, He J, Mi J, et al. Temporal relationship between inflammation and insulin resistance and their joint effect on hyperglycemia: the Bogalusa Heart Study. Cardiovasc Diabetol 2019;18.

35. Gala TD, Herder C, Rutters F, Carstensen-Kirberg M, Huth C, Stehouwer CDA, et al. Association of changes in inflammation with variation in glycaemia, insulin resistance and secretion based on the KORA study. Diabetes-Metab Res 2018;34.

36. Park K, Steffes M, Lee DH, Himes JH, Jacobs DR. Association of inflammation with worsening HOMA-insulin resistance. Diabetologia 2009;52: 2337–2344.

37. Herder C, Færch K, Carstensen-Kirberg M, Lowe GD, Haapakoski R, Witte DR, et al. Biomarkers of subclinical inflammation and increases in glycaemia, insulin resistance and beta-cell function in non-diabetic individuals: the Whitehall II study. Eur J Endocrinol 2016;175: 367–377.

38. Holman RR, Thorne KI, Farmer AJ, Davies MJ, Keenan JF, Paul S, et al. Addition of biphasic, prandial, or basal insulin to oral therapy in type 2 diabetes. N Engl J Med 2007;357: 1716–1730.

39. Intensive blood-glucose control with sulphonylureas or insulin compared with conventional treatment and risk of complications in patients with type 2 diabetes (UKPDS 33). UK Prospective Diabetes Study (UKPDS) Group. Lancet 1998;352: 837–853.

40. O’Hara L, Taylor J. What’s wrong with the’war on obesity’? A narrative review of the weight-centered health paradigm and development of the 3C framework to build critical competency for a paradigm shift. SAGE Open 2018.

41. Almuwaqqat Z, Hui Q, Liu C, Zhou JJ, Voight BF, Ho YL, et al. Long-Term Body Mass Index Variability and Adverse Cardiovascular Outcomes. JAMA Netw Open 2024;7: e243062.

42. Freeman L. A Matter of Justice: “Fat” Is Not Necessarily a Bad Word. Hastings Cent Rep 2020;50: 11–16.

43. Thille P, Friedman M, Setchell J. Weight-related stigma and health policy. CMAJ 2017;189: E223–E224.

44. Hunger JM, Smith JP, Tomiyama AJ. An evidence-based rational for adopting weight-inclusive health policy. Soc Issues Policy Rev 2020;14: 73–107.

