## Supplement for "Associations of body fat and inflammation with non-communicable chronic diseases and mortality: A prospective cohort study"

**Supplement legend**

Supplement Figure S1. Associations of body fat measures with cardiovascular disease

Supplement Figure S2. Associations of body fat measures with cancer

Supplement Figure S3. Associations of body fat measures with diabetes

Supplement Figure S4. Associations of body fat measures with asthma

Supplement Figure S5. Associations of body fat measures with gallbladder disease

Supplement Figure S6. Associations of body fat measures with back pain

Supplement Figure S7. Associations of body fat measures with osteoarthritis

Supplement Table S1. Baseline demographics and clinical characteristics by body mass index

Supplement Table S2. Associations of body fat with mortality and other clinical outcomes by sex, HR (95% CI) – primary analysis – $95^{th}$%ile vs $5^{th}$%ile

Supplement Table S3. Associations of body fat with mortality and other clinical outcomes by sex, HR (95% CI) – covariate analyses – $95^{th}$%ile vs $5^{th}$%ile

Supplement Table S4. Associations of body fat with clinical outcomes, with mortality modelled as a competing risk by sex, SHR (95% CI) – $95^{th}$%ile vs $5^{th}$%ile

Supplement Table S5. Associations of BMI or $fat_{BIA}$ and WHR with mortality and other clinical outcomes by sex, HR (95% CI) – $95^{th}$%ile vs $5^{th}$%ile

**Supplement Figure S1. Associations of body fat measures with cardiovascular disease**

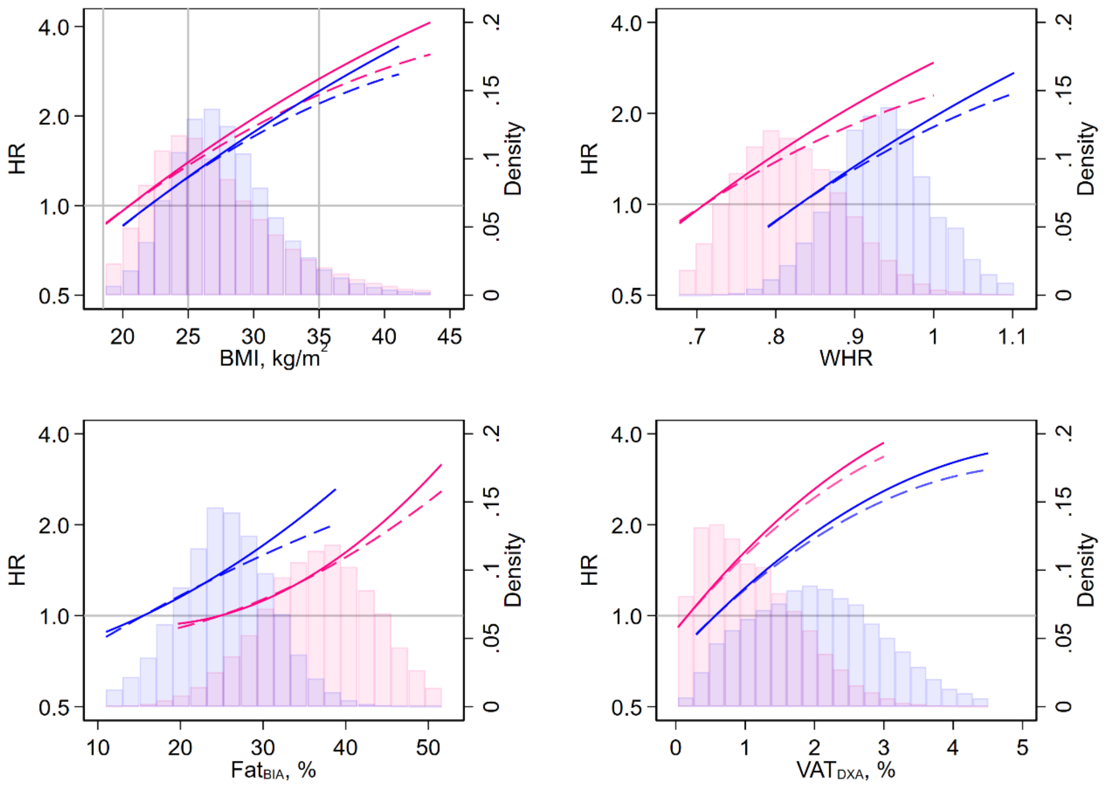


BIA bioimpedance analysis, BMI body mass index, DXA dual-energy X-ray absorptiometry, HR hazard ratio, lnCRP natural logarithm of C-reactive protein, VAT visceral adipose tissue, WHR waist:hip ratio

Results from four models are presented above (4 measures of body fat). Plots show hazard ratios plotted against BMI (top-left), WHR (top-right), $fat_{BIA}$ (bottom-left), and $VAT_{DXA}$ (bottom-right). Curves in pink represent females and curves in blue represent males. Solid lines represent HRs adjusted for age, sex, and lnCRP; and dashed lines represent fully adjusted HRs. The range of body fat represent the $1^{st}$%ile to the $99^{th}$%ile within sex. Histograms underlaying the HR plots show the distribution of the various measures of body fat by sex (pink for females, blue for males, and purple for overlapping distributions).

**Supplement Figure S2. Associations of body fat measures with cancer**

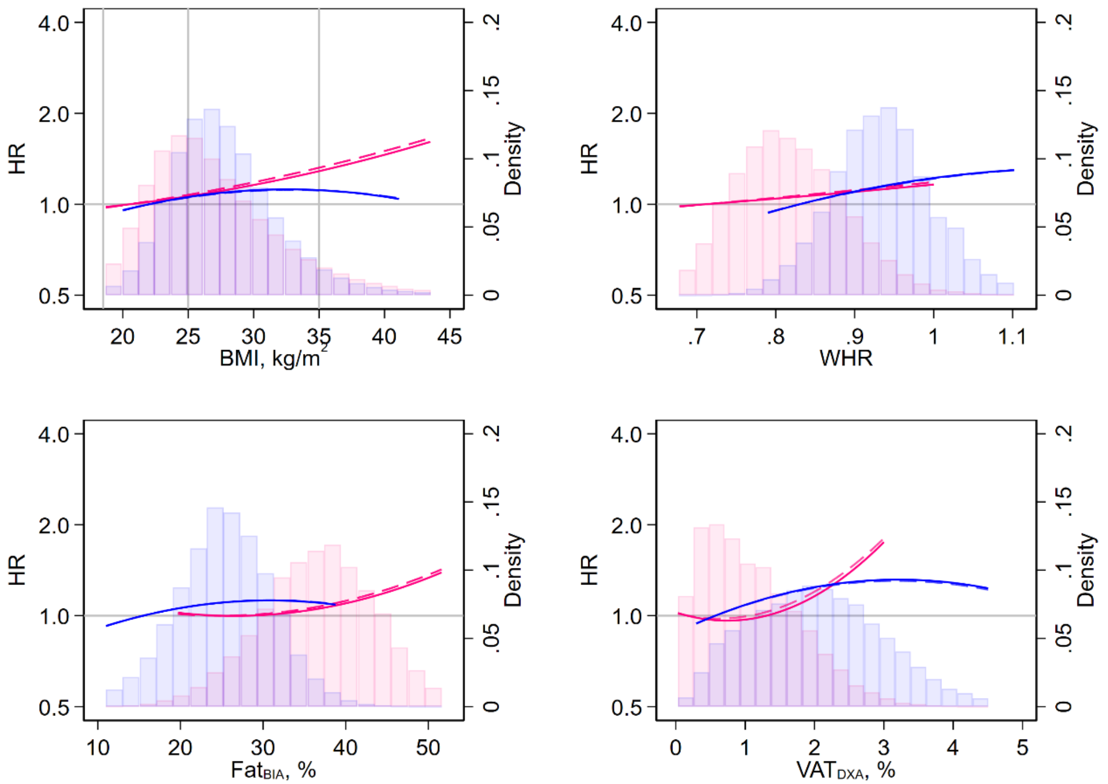


BIA bioimpedance analysis, BMI body mass index, DXA dual-energy X-ray absorptiometry, HR hazard ratio, lnCRP natural logarithm of C-reactive protein, VAT visceral adipose tissue, WHR waist:hip ratio

Results from four models are presented above (4 measures of body fat). Plots show hazard ratios plotted against BMI (top-left), WHR (top-right), $fat_{BIA}$ (bottom-left), and $VAT_{DXA}$ (bottom-right). Curves in pink represent females and curves in blue represent males. Solid lines represent HRs adjusted for age, sex, and lnCRP; and dashed lines represent fully adjusted HRs. The range of body fat represent the $1^{st}$%ile to the $99^{th}$%ile within sex. Histograms underlaying the HR plots show the distribution of the various measures of body fat by sex (pink for females, blue for males, and purple for overlapping distributions).

**Supplement Figure S3. Associations of body fat measures with diabetes**

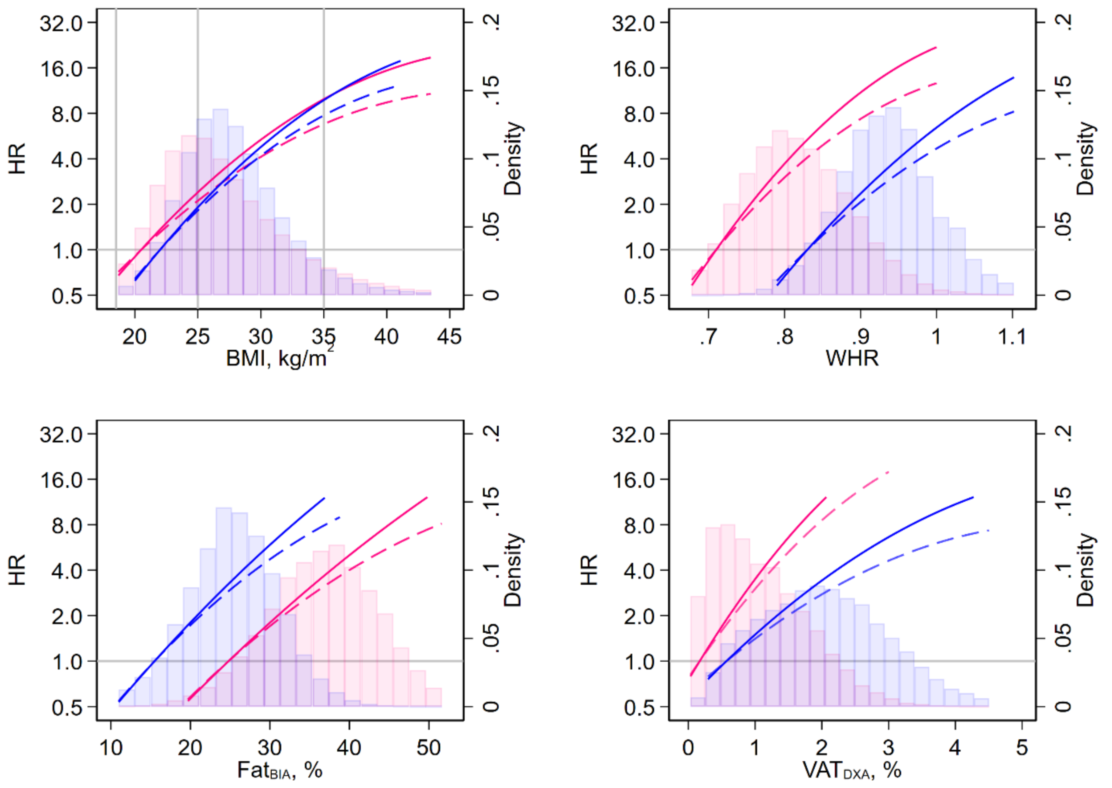


BIA bioimpedance analysis, BMI body mass index, DXA dual-energy X-ray absorptiometry, HR hazard ratio, lnCRP natural logarithm of C-reactive protein, VAT visceral adipose tissue, WHR waist:hip ratio

Results from four models are presented above (4 measures of body fat). Plots show hazard ratios plotted against BMI (top-left), WHR (top-right), $fat_{BIA}$ (bottom-left), and $VAT_{DXA}$ (bottom-right). Curves in pink represent females and curves in blue represent males. Solid lines represent HRs adjusted for age, sex, and lnCRP; and dashed lines represent fully adjusted HRs. The range of body fat represent the $1^{st}$%ile to the $99^{th}$%ile within sex. Histograms underlaying the HR plots show the distribution of the various measures of body fat by sex (pink for females, blue for males, and purple for overlapping distributions).

**Supplement Figure S4. Associations of body fat measures with asthma**

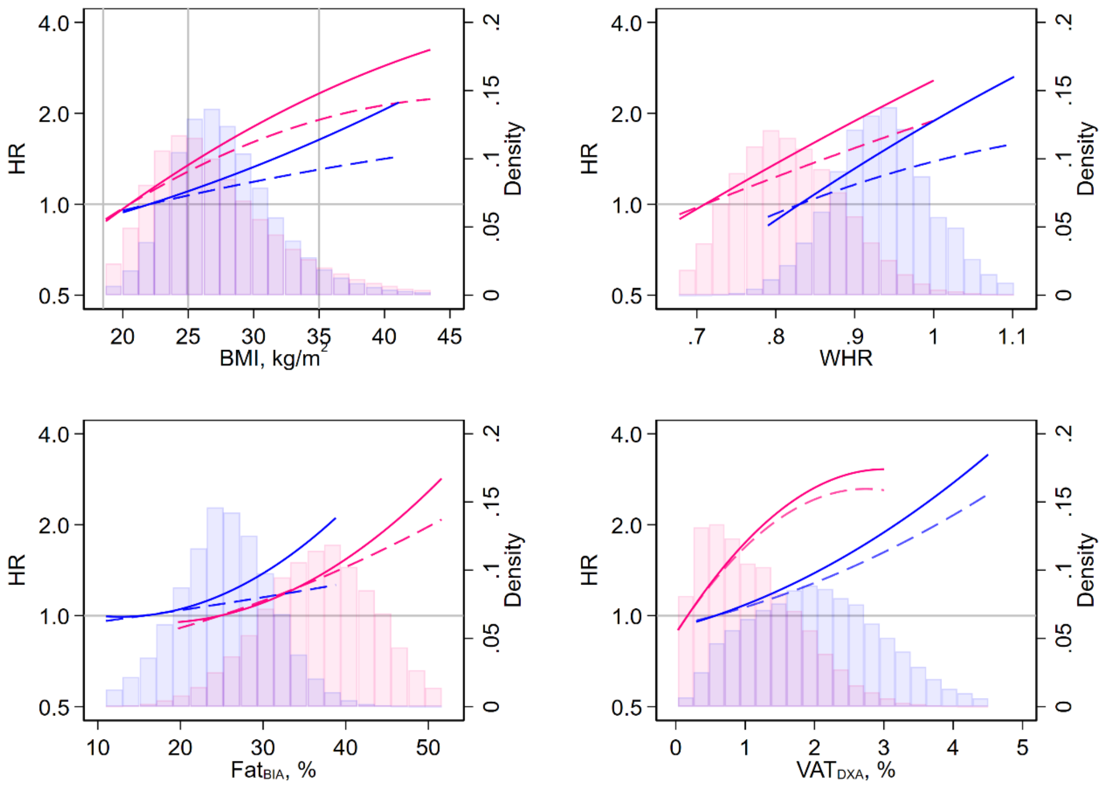


BIA bioimpedance analysis, BMI body mass index, DXA dual-energy X-ray absorptiometry, HR hazard ratio, lnCRP natural logarithm of C-reactive protein, VAT visceral adipose tissue, WHR waist:hip ratio

Results from four models are presented above (4 measures of body fat). Plots show hazard ratios plotted against BMI (top-left), WHR (top-right), $fat_{BIA}$ (bottom-left), and $VAT_{DXA}$ (bottom-right). Curves in pink represent females and curves in blue represent males. Solid lines represent HRs adjusted for age, sex, and lnCRP; and dashed lines represent fully adjusted HRs. The range of body fat represent the $1^{st}$%ile to the $99^{th}$%ile within sex. Histograms underlaying the HR plots show the distribution of the various measures of body fat by sex (pink for females, blue for males, and purple for overlapping distributions).

**Supplement Figure S5. Associations of body fat measures with gallbladder disease**

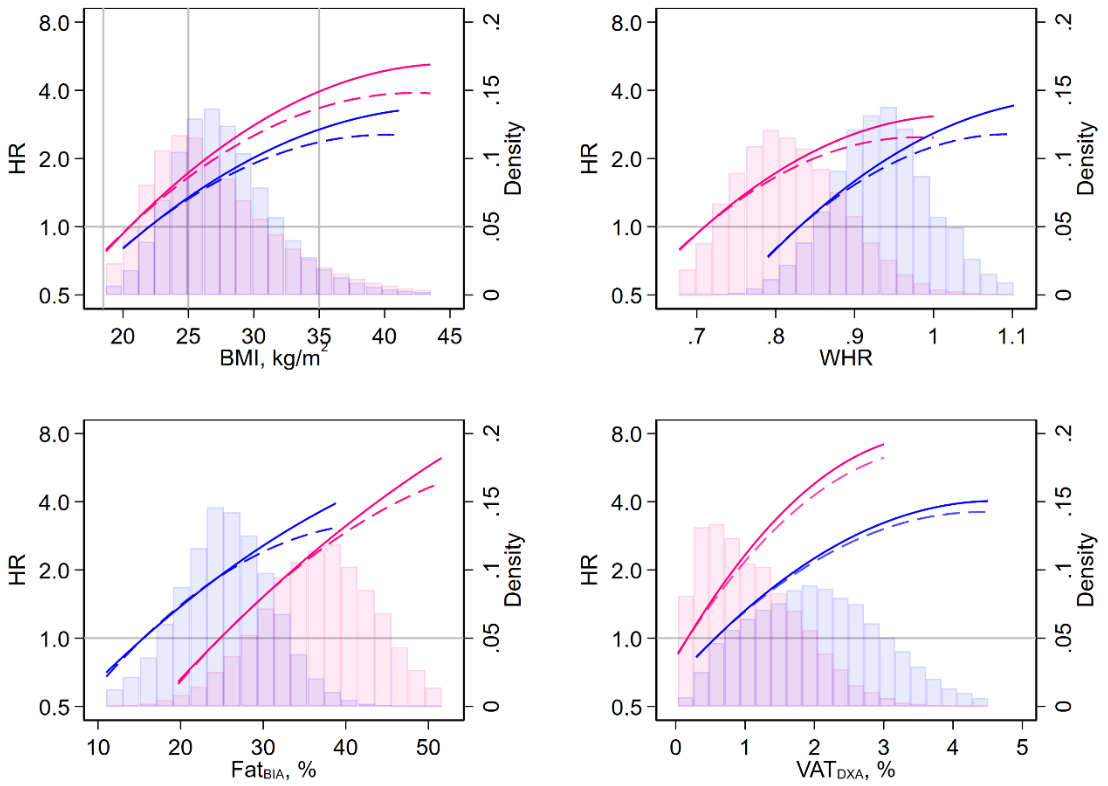


BIA bioimpedance analysis, BMI body mass index, DXA dual-energy X-ray absorptiometry, HR hazard ratio, lnCRP natural logarithm of C-reactive protein, VAT visceral adipose tissue, WHR waist:hip ratio

Results from four models are presented above (4 measures of body fat). Plots show hazard ratios plotted against BMI (top-left), WHR (top-right), $fat_{BIA}$ (bottom-left), and $VAT_{DXA}$ (bottom-right). Curves in pink represent females and curves in blue represent males. Solid lines represent HRs adjusted for age, sex, and lnCRP; and dashed lines represent fully adjusted HRs. The range of body fat represent the $1^{st}$%ile to the $99^{th}$%ile within sex. Histograms underlaying the HR plots show the distribution of the various measures of body fat by sex (pink for females, blue for males, and purple for overlapping distributions).

**Supplement Figure S6. Associations of body fat measures with back pain**

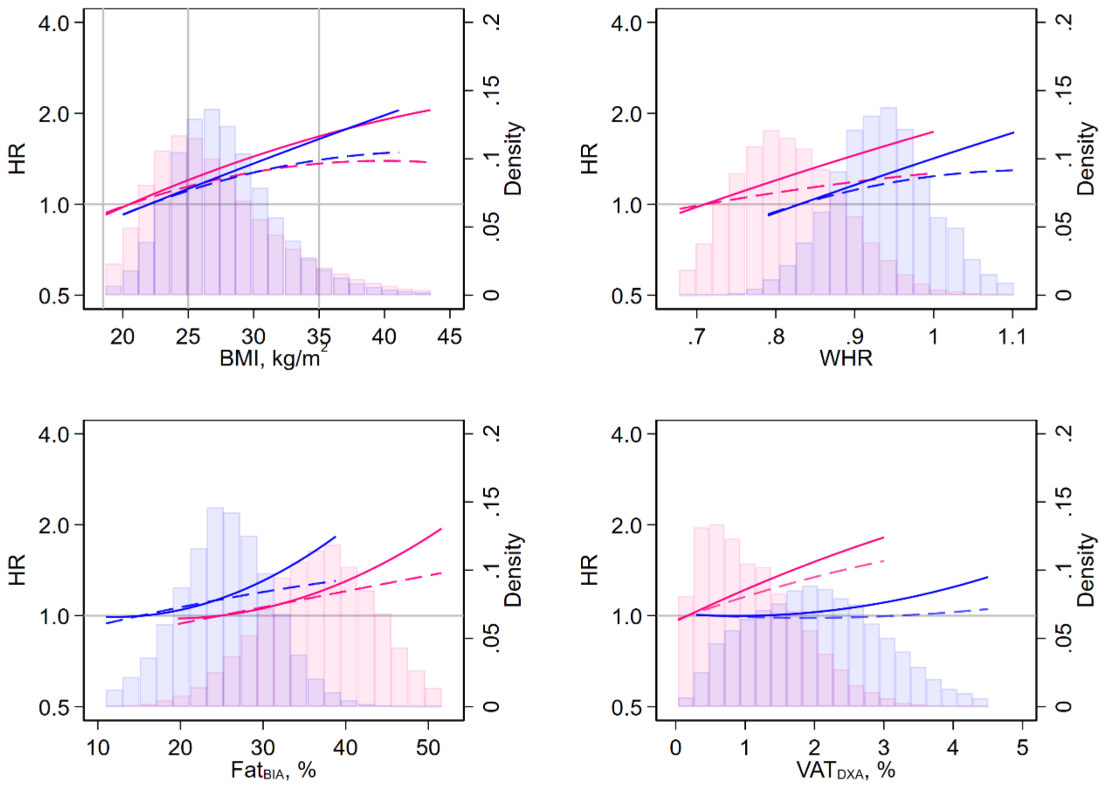


BIA bioimpedance analysis, BMI body mass index, DXA dual-energy X-ray absorptiometry, HR hazard ratio, lnCRP natural logarithm of C-reactive protein, VAT visceral adipose tissue, WHR waist:hip ratio

Results from four models are presented above (4 measures of body fat). Plots show hazard ratios plotted against BMI (top-left), WHR (top-right), $fat_{BIA}$ (bottom-left), and $VAT_{DXA}$ (bottom-right). Curves in pink represent females and curves in blue represent males. Solid lines represent HRs adjusted for age, sex, and lnCRP; and dashed lines represent fully adjusted HRs. The range of body fat represent the $1^{st}$%ile to the $99^{th}$%ile within sex. Histograms underlaying the HR plots show the distribution of the various measures of body fat by sex (pink for females, blue for males, and purple for overlapping distributions).

**Supplement Figure S7. Associations of body fat measures with osteoarthritis**

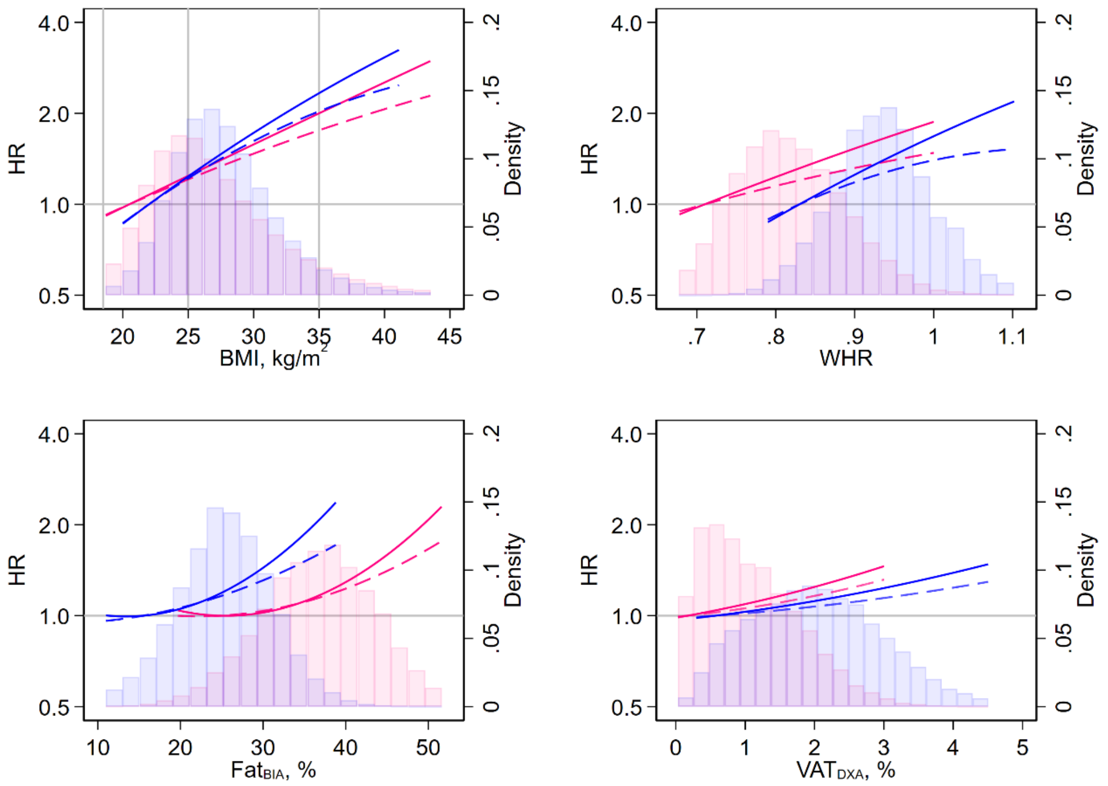


BIA bioimpedance analysis, BMI body mass index, DXA dual-energy X-ray absorptiometry, HR hazard ratio, lnCRP natural logarithm of C-reactive protein, VAT visceral adipose tissue, WHR waist:hip ratio

Results from four models are presented above (4 measures of body fat). Plots show hazard ratios plotted against BMI (top-left), WHR (top-right), $fat_{BIA}$ (bottom-left), and $VAT_{DXA}$ (bottom-right). Curves in pink represent females and curves in blue represent males. Solid lines represent HRs adjusted for age, sex, and lnCRP; and dashed lines represent fully adjusted HRs. The range of body fat represent the $1^{st}$%ile to the $99^{th}$%ile within sex. Histograms underlaying the HR plots show the distribution of the various measures of body fat by sex (pink for females, blue for males, and purple for overlapping distributions).

**Supplement Table S1. Baseline demographics and clinical characteristics by body mass index**

| Characteristic | <18.5 kg/m$^2$ | 18.5-24.9 kg/m$^2$ | 25.0-34.9 kg/m$^2$ | ≥35.0 kg/m$^2$ | P | Missing, % |
|---|---|---|---|---|---|---|
| N | 2,625 | 162,281 | 299,494 | 34,675 | - | 0.2 |
| Age, y | 56 [49,62] | 57 [49,63] | 58 [51,64] | 57 [50,63] | <0.001 | 0.0 |
| Male | 547 (20.8) | 56,700 (34.9) | 156,998 (52.4) | 13,104 (37.8) | <0.001 | 0.0 |
| Townsend deprivation index | -1.5 [-3.4,1.8] | -2.3 [-3.8,0.2] | -2.2 [-3.6,0.5] | -1.1 [-3.1,2.1] | <0.001 | 0.1 |
| Rural residence | 289 (11.1) | 23,643 (14.7) | 40,763 (13.8) | 3,685 (10.7) | <0.001 | 1.0 |
| Household income, £ | | | | | <0.001 | 15.3 |
| <18,000 | 669 (31.4) | 27,174 (19.7) | 59,167 (23.3) | 9,282 (32.3) | | |
| 18,000-30,999 | 510 (24.0) | 33,881 (24.5) | 65,579 (28.8) | 7,674 (26.7) | | |
| 31,000-51,999 | 495 (23.3) | 36,707 (26.6) | 66,177 (26.1) | 6,933 (24.1) | | |
| 52,000-100,000 | 339 (15.9) | 31,130 (22.5) | 50,386 (19.8) | 4,085 (14.2) | | |
| >100,000 | 116 (5.5) | 9,317 (6.7) | 12,640 (5.0) | 770 (2.7) | | |
| Employment | | | | | <0.001 | 1.3 |
| Yes | 1,338 (51.9) | 97,176 (60.7) | 168,939 (57.2) | 18,356 (53.6) | | |
| Retired | 770 (29.8) | 49,968 (31.2) | 104,237 (35.3) | 11,064 (32.3) | | |
| Unpaid/voluntary work | 195 (7.6) | 3,736 (2.3) | 9,964 (3.4) | 2,933 (8.6) | | |
| Homemaker | 56 (2.2) | 1,726 (1.1) | 1,917 (0.7) | 262 (0.8) | | |
| Disability | 140 (5.4) | 5,259 (3.3) | 5,721 (1.9) | 849 (2.5) | | |
| No | 81 (3.1) | 2,211 (1.4) | 4,779 (1.6) | 764 (2.2) | | |
| Post-secondary education | 1,620 (74.4) | 103,339 (74.2) | 172,066 (72.4) | 17,998 (69.4) | <0.001 | 18.9 |
| Non-white ethnicity | 166 (6.4) | 7,884 (4.9) | 16,169 (5.4) | 2,227 (6.5) | <0.001 | 0.5 |
| Government disability benefit | | | | | <0.001 | 1.0 |
| Living allowance | 210 (8.1) | 4,377 (2.7) | 13,175 (4.5) | 4,155 (12.2) | | |
| Attendance allowance | 15 (0.6) | 330 (0.2) | 916 (0.3) | 219 (0.6) | | |
| Blue badge | 17 (0.7) | 870 (0.5) | 3,655 (1.2) | 1,106 (3.2) | | |
| International physical activity questionnaire | | | | | <0.001 | 23.2 |
| High | 792 (40.7) | 57,341 (44.8) | 91,587 (40.0) | 6,767 (27.8) | | |
| Moderate | 832 (42.3) | 52,250 (40.8) | 93,230 (40.7) | 9,580 (39.4) | | |
| Low | 330 (17.0) | 18,374 (14.4) | 44,175 (19.3) | 7,959 (32.8) | | |
| Walking pace | | | | | <0.001 | 0.8 |
| Brisk | 1,417 (54.9) | 85,728 (53.1) | 102,384 (34.4) | 3,886 (11.4) | | |

| Characteristic | <18.5 kg/m$^2$ | 18.5-24.9 kg/m$^2$ | 25.0-34.9 kg/m$^2$ | ≥35.0 kg/m$^2$ | P | Missing, % |
|---|---|---|---|---|---|---|
| Steady average | 951 (36.8) | 69,614 (43.1) | 170,673 (57.4) | 20,404 (60.1) | | |
| Slow | 215 (8.3) | 6,093 (3.8) | 24,259 (8.2) | 9,679 (28.5) | | |
| Sleep duration, h | 7 [6,8] | 7 [7,8] | 7 [6,8] | 7 [6,8] | <0.001 | 0.8 |
| Insomnia | | | | | <0.001 | 0.2 |
| Usually | 798 (30.5) | 42,862 (26.5) | 84,152 (28.2) | 12,633 (36.5) | | |
| Sometimes | 1,244 (47.6) | 79,037 (48.8) | 142,087 (47.6) | 15,137 (43.8) | | |
| Never/rarely | 574 (21.9) | 40,089 (24.8) | 72,556 (24.3) | 6,808 (19.7) | | |
| Smoker | | | | | <0.001 | 0.5 |
| Current [pack-years] | 598 (22.9) [26.4] | 18,350 (11.4) [23.3] | 30,310 (10.2) [25.3] | 3,219 (9.4) [27.4] | <0.001 | |
| Previous [pack-years] | 540 (20.7) [12.0] | 47,763 (29.6) [12.0] | 110,486 (37.1) [18.3] | 13,350 (38.8) [23.3] | <0.001 | |
| Never | 1,473 (56.4) | 95,539 (59.1) | 157,073 (52.7) | 17,846 (51.9) | | |
| Alcohol consumption | | | | | <0.001 | 0.2 |
| Daily or almost daily | 580 (22.2) | 36,640 (22.6) | 60,375 (20.2) | 3,686 (10.7) | | |
| 3-4 times a week | 482 (18.5) | 39,775 (24.6) | 69,603 (23.3) | 5,062 (14.6) | | |
| 1-2 times a week | 487 (18.7) | 40,842 (25.2) | 78,631 (26.3) | 8,673 (25.1) | | |
| 1-3 times a month | 255 (9.8) | 16,738 (10.3) | 33,147 (11.1) | 5,420 (15.7) | | |
| Special occasions only | 401 (15.4) | 16,178 (10.0) | 33,759 (11.3) | 7,201 (20.8) | | |
| Former drinker | 192 (7.4) | 5,211 (3.2) | 10,371 (3.5) | 2,093 (6.1) | | |
| Never | 213 (8.2) | 6,588 (4.1) | 12,953 (4.3) | 2,431 (7.0) | | |
| Raw vegetable consumption | 2 [1,3] | 2 [1,3] | 2 [1,3] | 2 [1,3] | <0.001 | 1.5 |
| Fresh fruit consumption | 2 [1,3] | 2 [1,3] | 2 [1,3] | 2 [1,3] | <0.001 | 0.6 |
| Friends & family visits | | | | | <0.001 | 1.5 |
| Almost daily | 292 (11.4) | 16,415 (10.3) | 35,465 (12.0) | 5,220 (15.3) | | |
| 2-4 times a week | 678 (26.6) | 48,012 (30.0) | 91,802 (31.1) | 10,744 (31.6) | | |
| About once a week | 918 (35.9) | 59,169 (37.0) | 104,626 (35.5) | 10,770 (31.6) | | |
| About once a month | 369 (14.5) | 23,029 (14.4) | 38,571 (13.1) | 4,102 (12.1) | | |
| Once every few months | 221 (8.7) | 10,917 (6.8) | 19,460 (6.6) | 2,287 (6.7) | | |
| Never or almost never | 60 (2.4) | 2,064 (1.3) | 4,445 (1.5) | 743 (2.2) | | |
| No friends/family outside household | 16 (0.6) | 343 (0.2) | 740 (0.3) | 176 (0.5) | | |
| Leisure/social activities | | | | | <0.001 | 0.5 |

| Characteristic | <18.5 kg/m$^2$ | 18.5-24.9 kg/m$^2$ | 25.0-34.9 kg/m$^2$ | ≥35.0 kg/m$^2$ | P | Missing, % |
|---|---|---|---|---|---|---|
| Religious group | 462 (17.7) | 26,008 (16.1) | 45,865 (15.4) | 5,484 (15.9) | | |
| Pub or social club | 393 (15.1) | 33,271 (20.6) | 77,635 (26.1) | 7,547 (21.9) | | |
| Sports club or gym | 391 (15.0) | 34,312 (21.2) | 50,799 (17.1) | 4,184 (12.1) | | |
| Adult education class | 129 (5.0) | 5,610 (3.5) | 7,704 (2.6) | 1,017 (3.0) | | |
| Other group activity | 284 (10.9) | 15,505 (9.6) | 25,761 (8.6) | 3,245 (9.4) | | |
| None of the above | 948 (36.4) | 46,895 (29.0) | 90,245 (30.3) | 90,245 (30.3) | | |
| Able to confide | | | | | <0.001 | 3.7 |
| Almost daily | 1,185 (47.3) | 83,673 (53.4) | 155,514 (53.9) | 16,871 (50.7) | | |
| 2-4 times a week | 287 (11.4) | 16,596 (10.6) | 26,500 (9.2) | 3,138 (9.4) | | |
| About once a week | 340 (13.6) | 18,635 (11.9) | 30,403 (10.6) | 3,633 (10.9) | | |
| About once a month | 149 (5.9) | 8,931 (5.7) | 14,827 (5.1) | 1,798 (5.4) | | |
| Once every few months | 180 (7.2) | 8,841 (5.6) | 16,306 (5.7) | 1,854 (5.6) | | |
| Never or almost never | 367 (14.6) | 20,111 (12.8) | 44,753 (15.5) | 6,000 (18.0) | | |
| Comorbidities | | | | | | 0.0 |
| Cardiovascular disease | 358 (13.6) | 26,921 (16.6) | 97,867 (32.7) | 18,395 (53.0) | <0.001 | |
| Myocardial infarction | 33 (1.3) | 1,918 (1.2) | 8,390 (2.8) | 1,375 (4.0) | <0.001 | |
| Heart failure | 11 (0.4) | 425 (0.3) | 1,804 (0.6) | 453 (1.3) | <0.001 | |
| Hypertension | 297 (11.3) | 24,403 (15.0) | 91,349 (30.5) | 17,667 (51.0) | <0.001 | |
| Pulmonary embolism | 20 (0.8) | 895 (0.6) | 2,789 (0.9) | 614 (1.8) | <0.001 | |
| Stroke | 35 (1.3) | 1,627 (1.0) | 4,930 (1.6) | 862 (2.5) | <0.001 | |
| Cancer | 84 (3.2) | 5,025 (3.1) | 9,667 (3.2) | 1,179 (3.4) | 0.01 | |
| Diabetes | 33 (1.3) | 2,951 (1.8) | 16,980 (5.7) | 6,016 (17.3) | <0.001 | |
| Asthma | 265 (10.1) | 16,102 (9.9) | 33,968 (11.3) | 5,861 (16.9) | <0.001 | |
| Gallbladder disease | 43 (1.6) | 3,340 (2.1) | 11,328 (3.8) | 2,805 (8.1) | <0.001 | |
| Back pain | 197 (7.5) | 13,157 (8.1) | 29,776 (9.9) | 4,294 (12.4) | <0.001 | |
| Osteoarthritis | 155 (5.9) | 11,675 (7.2) | 33,485 (11.2) | 6,742 (19.4) | <0.001 | |
| Hand grip strength, kg | 26 [20,30] | 30 [24,38] | 32 [24,42] | 30 [22,38] | <0.001 | 0.1 |
| Forced expiratory volume in 1s, L | 2.5 [2.1,2.9] | 2.7 [2.3,3.3] | 2.8 [2.3,3.4] | 2.4 [2.0,3.0] | <0.001 | 9.4 |
| Neutrophil:lymphocyte ratio | 2.3 [1.8,3.1] | 2.2 [1.7,2.8] | 2.1 [1.7,2.7] | 2.2 [1.7,2.8] | <0.001 | 4.8 |
| Platelet:lymphocyte ratio | 149 [115,194] | 139 [111,176] | 130 [104,163] | 123 [97,155] | <0.001 | 4.8 |
| Red cell distribution width, % | 13.4 [12.9,14.1] | 13.3 [12.9,13.8] | 13.3 [12.9,13.9] | 13.6 [13.1,14.2] | <0.001 | 4.6 |
| Apolipoprotein B, g/L | 0.9 [0.8,1.0] | 1.0 [0.8,1.1] | 1.0 [0.9,1.2] | 1.0 [0.9,1.2] | <0.001 | 6.8 |
| C-reactive protein, mg/L | 0.4 [0.2,1.0] | 0.8 [0.4,1.6] | 1.6 [0.8,3.0] | 3.8 [2.1,7.0] | <0.001 | 6.6 |

| Characteristic | <18.5 kg/m$^2$ | 18.5-24.9 kg/m$^2$ | 25.0-34.9 kg/m$^2$ | ≥35.0 kg/m$^2$ | P | Missing, % |
|---|---|---|---|---|---|---|
| Glycated hemoglobin, % | 5.3 [5.1,5.6] | 5.3 [5.1,5.5] | 5.4 [5.2,5.6] | 5.6 [5.3,6.0] | <0.001 | 7.0 |
| Physical measures | | | | | | |
| Body mass index, kg/m$^2$ | 17.9 [17.3,18.2] | 23.1 [21.8,24.1] | 28.2 [26.5,30.4] | 37.7 [36.1,40.4] | <0.001 | 0.2 |
| Waist:hip ratio | 0.76 [0.73,0.80] | 0.81 [0.76,0.88] | 0.90 [0.83,0.95] | 0.92 [0.85,1.00] | <0.001 | <0.1 |
| Bioimpedance analysis | | | | | | |
| Body fat, % | 19.3 [14.9,22.5] | 27.7 [21.5,32.5] | 32.4 [26.3,39.2] | 45.0 [36.8,48.4] | <0.001 | 1.7 |
| Dual-energy X-ray absorptiometry | | | | | | |
| Tissue fat, % | 24.5 [19.8,28.9] | 31.5 [25.9,36.8] | 36.1 [30.8,42.3] | 46.9 [41.9,51.4] | <0.001 | 90.5 |
| Visceral adipose tissue, % | 0.36 [0.22,0.58] | 0.79 [0.45,1.32] | 1.83 [1.23,2.49] | 2.36 [1.73,3.13] | <0.001 | 92.0 |
| Abdominal magnetic resonance imaging | | | | | | |
| Adipose tissue index, L/m$^2$ | 1.22 [0.90,1.61] | 2.55 [1.91,3.23] | 4.09 [3.29,5.03] | 7.25 [6.10,8.38] | <0.001 | 92.0 |
| Subcutaneous adipose tissue, % | 25.6 [21.9,29.8] | 31.1 [27.5,35.1] | 32.6 [28.6,37.2] | 37.9 [34.8,41.5] | <0.001 | 98.3 |
| Visceral adipose tissue, % | 7.5 [5.7,10.7] | 11.7 [8.3,17.4] | 18.7 [12.5,25.3] | 15.1 [11.9,22.0] | <0.001 | 98.3 |

£ UK pound sterling

N (%) or median [inter-quartile range] are reported. Differences across categories of body mass index are tested using the Chi-square and Kruskal-Wallis tests, as appropriate. All categorical variables were mutually exclusive. There were 1,032 participants included in the study without a measurement of body mass index and thus are not described in this table.

**Supplement Table S2. Associations of body fat with mortality and other clinical outcomes by sex, HR (95% CI) – primary analysis – 95th%ile vs 5th%ile**

| Outcome | Events (%) | BMI, kg/m² | WHR | $Fat_{BIA}$, % | $Fat_{DXA}$, % | $VAT_{DXA}$, % | $Index_{MRI}$, L/m² |
|---|---|---|---|---|---|---|---|
| *Females* | | | | | | | |
| N<br>5th%ile-95th%ile | ≤272,163<br>- | 271,726<br>20.5-37.0 | 272,058<br>0.71-0.94 | 268,064<br>24.8-47.6 | 24,667<br>26.2-50.5 | 20,625<br>0.17-2.38 | 20,774<br>1.54-7.24 |
| Mortality | 18,140 (6.7) | *0.98 (0.93,1.03)* | 1.69 (1.61,1.77) | **0.93 (0.89,0.98)** | *1.08 (0.74,1.58)* | *0.75 (0.49,1.13)* | *0.79 (0.52,1.20)* |
| Cardiovascular | 38,439 (18.7) | 2.99 (2.88,3.10) | 2.41 (2.32,2.49) | 2.44 (2.35,2.54) | 2.71 (2.38,3.10) | 3.05 (2.65,3.51) | 3.21 (2.76,3.72) |
| Myocardial infarction | 5,799 (2.2) | 1.81 (1.65,1.99) | 2.73 (2.48,3.00) | 1.67 (1.52,1.84) | 2.63 (1.61,4.31) | 2.38 (1.48,3.81) | 2.75 (1.68,4.49) |
| Heart failure | 7,014 (2.6) | 2.87 (2.64,3.12) | 2.83 (2.62,3.05) | 2.11 (1.93,2.31) | 2.22 (1.42,3.47) | 1.87 (1.11,3.15) | 2.28 (1.33,3.91) |
| Hypertension | 34,039 (16.3) | 3.41 (3.28,3.55) | 2.62 (2.52,2.72) | 2.70 (2.59,2.81) | 2.89 (2.51,3.32) | 3.23 (2.78,3.74) | 3.57 (3.05,4.18) |
| Pulmonary embolism | 4,367 (1.6) | 3.08 (2.76,3.43) | 1.99 (1.79,2.22) | 2.82 (2.50,3.19) | 6.22 (3.30,11.72) | 3.69 (2.07,6.56) | 5.11 (2.81,9.32) |
| Stroke | 5,299 (2.0) | 1.24 (1.13,1.37) | 1.57 (1.44,1.72) | 1.15 (1.05,1.27) | *1.12 (0.71,1.78)* | *1.21 (0.75,1.97)* | *0.80 (0.47,1.34)* |
| Cancer | 15,949 (6.1) | 1.35 (1.28,1.43) | 1.12 (1.07,1.18) | 1.26 (1.19,1.33) | 1.93 (1.55,2.40) | 1.32 (1.06,1.64) | 1.58 (1.25,2.00) |
| Diabetes | 11,185 (4.3) | 12.18 (11.27,13.17) | 14.46 (13.20,15.83) | 10.09 (9.09,11.20) | 5.48 (3.77,7.98) | 16.32 (11.02,24.17) | 11.05 (7.49,16.30) |
| Asthma | 7,583 (3.2) | 2.55 (2.35,2.77) | 2.11 (1.96,2.28) | 2.24 (2.05,2.44) | 3.09 (2.20,4.34) | 2.89 (2.06,4.06) | 3.44 (2.38,4.97) |
| Gallbladder disease | 12,900 (5.0) | 4.39 (4.10,4.70) | 2.78 (2.60,2.97) | 5.01 (4.62,5.44) | 5.75 (4.28,7.73) | 5.82 (4.44,7.61) | 7.16 (5.35,9.59) |
| Back pain | 20,546 (8.4) | 1.77 (1.69,1.87) | 1.55 (1.48,1.63) | 1.66 (1.58,1.74) | 1.76 (1.48,2.10) | 1.62 (1.35,1.95) | 1.94 (1.59,2.36) |
| Osteoarthritis | 24,322 (10.2) | 2.20 (2.10,2.31) | 1.65 (1.58,1.72) | 1.81 (1.73,1.90) | 1.58 (1.37,1.83) | 1.32 (1.13,1.54) | 1.62 (1.38,1.91) |

| Outcome | Events (%) | BMI, kg/m$^2$ | WHR | $Fat_{BIA}$, % | $Fat_{DXA}$, % | $VAT_{DXA}$, % | $Index_{MRI}$, L/m$^2$ |
|---|---|---|---|---|---|---|---|
| *Males* | | | | | | | |
| N<br>5th%ile-95th%ile | ≤227,944<br>- | 227,349<br>22.0-35.4 | 227,856<br>0.83-1.05 | 223,716<br>15.5-34.7 | 23,080<br>18.7-40.8 | 19,431<br>0.57-3.70 | 19,298<br>1.40-5.99 |
| Mortality | 26,005 (11.4) | 1.11 (1.07,1.16) | 1.91 (1.83,1.99) | 1.17 (1.12,1.22) | *1.29 (0.98,1.70)* | *1.07 (0.78,1.49)* | *1.24 (0.92,1.67)* |
| Cardiovascular | 38,543 (25.6) | 2.50 (2.41,2.59) | 2.26 (2.18,2.35) | 2.13 (2.05,2.21) | 2.62 (2.32,2.96) | 3.05 (2.68,3.47) | 3.40 (2.97,3.88) |
| Myocardial infarction | 12,182 (5.6) | 1.77 (1.66,1.88) | 2.11 (1.97,2.26) | 1.71 (1.59,1.83) | 1.38 (1.08,1.76) | 1.69 (1.28,2.23) | 1.75 (1.33,2.29) |
| Heart failure | 11,524 (5.1) | 2.74 (2.57,2.92) | 3.11 (2.91,3.32) | 1.95 (1.83,2.09) | 1.91 (1.41,2.59) | 1.92 (1.36,2.72) | 2.70 (1.90,3.84) |
| Hypertension | 34,338 (21.9) | 2.94 (2.83,3.05) | 2.61 (2.51,2.71) | 2.49 (2.40,2.59) | 2.98 (2.62,3.40) | 3.37 (2.93,3.87) | 3.82 (3.31,4.41) |
| Pulmonary embolism | 4,642 (2.1) | 1.86 (1.68,2.05) | 1.85 (1.67,2.05) | 1.59 (1.44,1.76) | 2.88 (1.83,4.54) | 2.35 (1.48,3.74) | 3.43 (2.09,5.61) |
| Stroke | 6,897 (3.1) | 1.22 (1.13,1.33) | 1.54 (1.42,1.68) | 1.18 (1.08,1.28) | *1.20 (0.86,1.67)* | 1.72 (1.13,2.61) | *1.26 (0.85,1.85)* |
| Cancer | 17,139 (7.7) | 1.11 (1.05,1.17) | 1.26 (1.19,1.33) | 1.11 (1.06,1.18) | 1.55 (1.27,1.89) | 1.30 (1.06,1.61) | 1.48 (1.19,1.85) |
| Diabetes | 14,199 (6.7) | 10.38 (9.70,11.11) | 9.23 (8.54,9.97) | 9.68 (8.89,10.54) | 4.43 (3.40,5.76) | 9.59 (7.03,13.07) | 8.32 (6.24,11.11) |
| Asthma | 5,599 (2.7) | 1.67 (1.53,1.83) | 2.19 (2.00,2.41) | 1.69 (1.54,1.86) | 1.65 (1.21,2.27) | 2.44 (1.70,3.49) | 2.58 (1.77,3.76) |
| Gallbladder disease | 8,170 (3.7) | 2.75 (2.54,2.98) | 2.98 (2.74,3.26) | 3.26 (2.98,3.57) | 3.33 (2.40,4.63) | 3.76 (2.68,5.28) | 3.44 (2.43,4.87) |
| Back pain | 14,954 (7.2) | 1.67 (1.58,1.77) | 1.64 (1.55,1.73) | 1.54 (1.45,1.63) | 1.27 (1.05,1.53) | *1.19 (0.98,1.46)* | 1.33 (1.08,1.63) |
| Osteoarthritis | 15,811 (7.6) | 2.40 (2.27,2.53) | 1.89 (1.79,2.00) | 1.83 (1.73,1.94) | 1.52 (1.28,1.81) | 1.34 (1.10,1.62) | 1.60 (1.32,1.94) |

BIA bioimpedance analysis, BMI body mass index, CI confidence interval, DXA dual-energy X-ray absorptiometry, HR hazard ratio, lnCRP natural logarithm of c-reactive protein, MRI magnetic resonance imaging, WHR waist:hip ratio, VAT visceral adipose tissue

The models are adjusted for age, sex, lnCRP and a measure of body fat (linear and quadratic terms). Sex is interacted with the measures of body fat. Significant negative associations are in blue font and bolded. Non-significant associations are in italics.

**Supplement Table S3. Associations of body fat with mortality and other clinical outcomes by sex, HR (95% CI) – covariate analyses – 95th%ile vs 5th%ile**

| Outcome | Females | | | Males | | |
|---|---|---|---|---|---|---|
| | BMI | WHR | $Fat_{BIA}$ | BMI | WHR | $Fat_{BIA}$ |
| *Fully adjusted* | | | | | | |
| Mortality | **0.80 (0.76,0.84)** | 1.21 (1.16,1.28) | **0.80 (0.76,0.84)** | **0.89 (0.85,0.93)** | 1.19 (1.14,1.24) | **0.89 (0.85,0.92)** |
| Cardiovascular | 2.58 (2.48,2.68) | 2.01 (1.94,2.08) | 2.14 (2.06,2.22) | 2.25 (2.17,2.33) | 1.83 (1.77,1.90) | 1.81 (1.74,1.88) |
| Cancer | 1.39 (1.31,1.47) | 1.14 (1.08,1.20) | 1.29 (1.22,1.37) | 1.11 (1.05,1.17) | 1.26 (1.19,1.33) | 1.12 (1.06,1.18) |
| Diabetes | 7.96 (7.35,8.62) | 9.14 (8.35,10.02) | 6.60 (5.94,7.32) | 8.14 (7.60,8.72) | 6.14 (5.67,6.64) | 6.84 (6.28,7.46) |
| Asthma | 2.01 (1.84,2.19) | 1.63 (1.51,1.76) | 1.82 (1.67,1.99) | 1.31 (1.20,1.44) | 1.48 (1.34,1.62) | 1.21 (1.10,1.33) |
| Gallbladder disease | 3.61 (3.37,3.87) | 2.39 (2.23,2.56) | 4.21 (3.88,4.58) | 2.40 (2.21,2.60) | 2.45 (2.24,2.67) | 2.82 (2.57,3.09) |
| Back pain | 1.38 (1.31,1.46) | 1.22 (1.16,1.28) | 1.32 (1.25,1.39) | 1.41 (1.33,1.49) | 1.27 (1.20,1.35) | 1.25 (1.18,1.32) |
| Osteoarthritis | 1.88 (1.79,1.97) | 1.38 (1.32,1.44) | 1.53 (1.46,1.60) | 2.07 (1.96,2.19) | 1.46 (1.38,1.55) | 1.50 (1.41,1.58) |
| *Age-sex adjusted* | | | | | | |
| Mortality | 1.44 (1.38,1.52) | 2.10 (2.00,2.20) | 1.37 (1.31,1.44) | 1.40 (1.35,1.46) | 2.33 (2.24,2.43) | 1.52 (1.46,1.58) |
| Cardiovascular | 3.76 (3.63,3.90) | 2.89 (2.79,2.99) | 3.15 (3.04,3.27) | 2.89 (2.80,3.00) | 2.70 (2.61,2.80) | 2.56 (2.47,2.65) |
| Cancer | 1.42 (1.35,1.50) | 1.17 (1.12,1.24) | 1.34 (1.27,1.41) | 1.15 (1.09,1.21) | 1.32 (1.25,1.38) | 1.16 (1.10,1.22) |
| Diabetes | 18.90 (17.53,20.38) | 21.43 (19.59,23.45) | 17.16 (15.49,19.02) | 13.55 (12.68,14.48) | 13.12 (12.15,14.16) | 13.73 (12.62,14.93) |
| Asthma | 3.20 (2.96,3.46) | 2.53 (2.35,2.72) | 2.86 (2.64,3.10) | 1.92 (1.76,2.10) | 2.59 (2.37,2.84) | 2.00 (1.83,2.19) |

| Outcome | Females | | | Males | | |
|---|---|---|---|---|---|---|
| | BMI | WHR | $Fat_{BIA}$ | BMI | WHR | $Fat_{BIA}$ |
| Gallbladder disease | 6.10 (5.73,6.50) | 3.74 (3.51,3.99) | 6.95 (6.43,7.51) | 3.36 (3.10,3.63) | 3.89 (3.57,4.24) | 4.07 (3.73,4.45) |
| Back pain | 2.02 (1.93,2.12) | 1.74 (1.67,1.82) | 1.91 (1.82,2.00) | 1.81 (1.72,1.92) | 1.82 (1.72,1.92) | 1.69 (1.60,1.79) |
| Osteoarthritis | 2.53 (2.42,2.64) | 1.90 (1.83,1.98) | 2.16 (2.07,2.25) | 2.61 (2.47,2.75) | 2.16 (2.04,2.28) | 2.06 (1.95,2.18) |
| *Adjusted for social determinants* | | | | | | |
| Mortality | 1.20 (1.14,1.26) | 1.76 (1.68,1.85) | 1.16 (1.11,1.22) | 1.22 (1.17,1.27) | 1.84 (1.77,1.92) | 1.28 (1.23,1.33) |
| Cardiovascular | 3.40 (3.29,3.53) | 2.63 (2.54,2.72) | 2.87 (2.77,2.98) | 2.77 (2.67,2.86) | 2.45 (2.37,2.54) | 2.37 (2.29,2.46) |
| Cancer | 1.45 (1.38,1.53) | 1.20 (1.14,1.26) | 1.36 (1.29,1.44) | 1.15 (1.09,1.21) | 1.32 (1.26,1.40) | 1.17 (1.11,1.23) |
| Diabetes | 15.13 (14.03,16.32) | 16.75 (15.30,18.33) | 13.91 (12.56,15.41) | 12.30 (11.52,13.14) | 10.86 (10.05,11.73) | 11.83 (10.88,12.87) |
| Asthma | 2.78 (2.57,3.01) | 2.18 (2.02,2.34) | 2.51 (2.31,2.72) | 1.77 (1.62,1.94) | 2.20 (2.00,2.41) | 1.77 (1.62,1.94) |
| Gallbladder disease | 5.49 (5.15,5.85) | 3.41 (3.20,3.64) | 6.30 (5.82,6.81) | 3.13 (2.89,3.38) | 3.45 (3.16,3.76) | 3.77 (3.45,4.12) |
| Back pain | 1.77 (1.69,1.86) | 1.52 (1.45,1.59) | 1.68 (1.60,1.76) | 1.69 (1.60,1.78) | 1.57 (1.48,1.66) | 1.51 (1.43,1.60) |
| Osteoarthritis | 2.31 (2.21,2.41) | 1.73 (1.66,1.81) | 1.98 (1.89,2.06) | 2.40 (2.27,2.53) | 1.89 (1.78,1.99) | 1.87 (1.77,1.97) |
| *Adjusted for behaviours* | | | | | | |
| Mortality | 1.34 (1.28,1.41) | 1.80 (1.72,1.89) | 1.30 (1.24,1.36) | 1.38 (1.33,1.44) | 2.03 (1.95,2.11) | 1.45 (1.39,1.51) |
| Cardiovascular | 3.59 (3.46,3.71) | 2.70 (2.61,2.80) | 3.03 (2.92,3.14) | 2.85 (2.75,2.95) | 2.55 (2.46,2.64) | 2.50 (2.41,2.59) |
| Cancer | 1.45 (1.38,1.53) | 1.17 (1.12,1.24) | 1.35 (1.28,1.43) | 1.15 (1.10,1.22) | 1.30 (1.23,1.37) | 1.15 (1.09,1.22) |
| Diabetes | 15.77 (14.62,17.01) | 18.25 (16.68,19.97) | 14.61 (13.19,16.19) | 12.76 (11.95,13.64) | 11.91 (11.02,12.86) | 13.06 (12.01,14.21) |

| Outcome | Females | | | Males | | |
|---|---|---|---|---|---|---|
| | BMI | WHR | $Fat_{BIA}$ | BMI | WHR | $Fat_{BIA}$ |
| Asthma | 2.99 (2.76,3.23) | 2.31 (2.15,2.49) | 2.71 (2.50,2.94) | 1.86 (1.70,2.03) | 2.40 (2.19,2.63) | 1.94 (1.77,2.12) |
| Gallbladder disease | 5.34 (5.01,5.69) | 3.36 (3.15,3.58) | 6.15 (5.69,6.65) | 3.17 (2.93,3.43) | 3.57 (3.27,3.89) | 3.83 (3.50,4.18) |
| Back pain | 1.88 (1.79,1.97) | 1.59 (1.52,1.67) | 1.80 (1.72,1.89) | 1.76 (1.67,1.86) | 1.70 (1.61,1.80) | 1.65 (1.56,1.74) |
| Osteoarthritis | 2.42 (2.32,2.53) | 1.80 (1.73,1.88) | 2.09 (2.00,2.18) | 2.51 (2.37,2.65) | 2.05 (1.94,2.16) | 2.00 (1.90,2.12) |
| *Adjusted for labs* | | | | | | |
| Mortality | *0.97 (0.92,1.01)* | 1.68 (1.61,1.77) | **0.95 (0.90,0.99)** | 1.10 (1.06,1.15) | 1.84 (1.76,1.92) | 1.19 (1.15,1.24) |
| Cardiovascular | 2.90 (2.79,3.01) | 2.35 (2.27,2.43) | 2.40 (2.31,2.49) | 2.43 (2.35,2.52) | 2.20 (2.12,2.28) | 2.09 (2.02,2.17) |
| Cancer | 1.35 (1.28,1.43) | 1.14 (1.08,1.20) | 1.27 (1.20,1.35) | 1.12 (1.06,1.18) | 1.27 (1.20,1.33) | 1.12 (1.06,1.18) |
| Diabetes | 11.78 (10.89,12.75) | 14.17 (12.93,15.52) | 9.87 (8.89,10.96) | 10.50 (9.80,11.24) | 9.03 (8.35,9.76) | 9.76 (8.96,10.65) |
| Asthma | 2.54 (2.33,2.76) | 2.14 (1.98,2.30) | 2.27 (2.09,2.48) | 1.66 (1.52,1.82) | 2.15 (1.96,2.36) | 1.70 (1.55,1.86) |
| Gallbladder disease | 4.35 (4.07,4.66) | 2.80 (2.62,2.99) | 5.07 (4.67,5.50) | 2.73 (2.52,2.95) | 2.93 (2.69,3.20) | 3.26 (2.98,3.57) |
| Back pain | 1.78 (1.69,1.87) | 1.57 (1.50,1.65) | 1.68 (1.60,1.77) | 1.67 (1.58,1.76) | 1.62 (1.53,1.71) | 1.54 (1.45,1.63) |
| Osteoarthritis | 2.20 (2.10,2.30) | 1.68 (1.61,1.75) | 1.84 (1.76,1.93) | 2.38 (2.25,2.51) | 1.87 (1.76,1.97) | 1.84 (1.74,1.94) |
| *Adjusted for comorbidities and other health related measures* | | | | | | |
| Mortality | *0.99 (0.94,1.04)* | 1.50 (1.43,1.57) | *0.99 (0.95,1.04)* | **0.94 (0.90,0.98)** | 1.43 (1.37,1.49) | *0.98 (0.94,1.02)* |
| Cardiovascular | 3.19 (3.08,3.31) | 2.50 (2.41,2.59) | 2.71 (2.61,2.81) | 2.51 (2.43,2.60) | 2.20 (2.13,2.28) | 2.10 (2.02,2.17) |
| Cancer | 1.42 (1.35,1.50) | 1.17 (1.11,1.23) | 1.33 (1.26,1.41) | 1.13 (1.08,1.20) | 1.31 (1.25,1.39) | 1.16 (1.10,1.22) |

| Outcome | Females | | | Males | | |
|---|---|---|---|---|---|---|
| | BMI | WHR | $Fat_{BIA}$ | BMI | WHR | $Fat_{BIA}$ |
| Diabetes | 12.90 (11.95,13.92) | 14.82 (13.54,16.23) | 11.79 (10.64,13.07) | 9.41 (8.80,10.07) | 8.11 (7.50,8.77) | 8.68 (7.97,9.44) |
| Asthma | 2.29 (2.11,2.48) | 1.86 (1.73,2.00) | 2.08 (1.92,2.26) | 1.40 (1.28,1.53) | 1.67 (1.52,1.83) | 1.31 (1.20,1.44) |
| Gallbladder disease | 5.17 (4.84,5.52) | 3.13 (2.93,3.34) | 5.94 (5.49,6.42) | 2.90 (2.70,3.15) | 3.09 (2.83,3.37) | 3.41 (3.11,3.73) |
| Back pain | 1.51 (1.44,1.59) | 1.35 (1.29,1.42) | 1.44 (1.37,1.51) | 1.44 (1.36,1.52) | 1.37 (1.29,1.45) | 1.29 (1.22,1.36) |
| Osteoarthritis | 2.06 (1.97,2.15) | 1.53 (1.47,1.60) | 1.72 (1.64,1.79) | 2.23 (2.11,2.36) | 1.66 (1.56,1.75) | 1.64 (1.55,1.73) |

BIA bioimpedance analysis, BMI body mass index, CI confidence interval, HR hazard ratio, IPAQ International Physical Activity Questionnaire, lnCRP natural logarithm of c-reactive protein, NLR neutrophil:lymphocyte ratio, PLR platelet:lymphocyte ratio, RDW red cell distribution width, WHR waist:hip ratio

Results from six models are presented above. The first model adjusted for all covariates listed in Table 1 except for IPAQ and walking pace which may be determined, at least in part, by a participant's body fat. The second model adjusted for age and sex but not lnCRP. The third model adjusted for age, sex, and social variables (Townsend deprivation index, rural residence, household income, employment, post-secondary education, non-white ethnicity, friends and family visits, leisure/social activities, able to confide). The fourth model adjusted for age, sex, and behaviours (alcohol consumption, fresh fruit consumption, sleep duration, smoker, raw vegetable consumption). The fifth model adjusted age, sex, and labs (lnCRP, apolipoprotein B, glycated hemoglobin, NLR, PLR, RDW - all quadratic terms except CRP). The sixth model adjusted for age, sex, and baseline comorbidities (cardiovascular disease, cancer, diabetes, asthma, osteoarthritis, gallbladder disease, chronic back pain) and other related health measures (forced expiratory volume in 1s, hand grip strength, government-accommodated disability, insomnia). Significant negative associations are in blue font and bolded. Non-significant associations are in italics.

**Supplement Table S4. Associations of body fat with clinical outcomes, with mortality modelled as a competing risk by sex, SHR (95% CI) – 95th%ile vs 5th%ile**

| Outcome | Females | | | Males | | |
|---|---|---|---|---|---|---|
| | BMI | WHR | $Fat_{BIA}$ | BMI | WHR | $Fat_{BIA}$ |
| Cardiovascular | 2.90 (2.80,3.01) | 2.38 (2.29,2.46) | 2.37 (2.28,2.46) | 3.02 (2.89,3.16) | 2.61 (2.44,2.78) | 3.07 (2.74,3.43) |
| Cancer | 1.07 (1.02,1.13) | *0.95 (0.91,1.003)* | *0.98 (0.93,1.03)* | *0.98 (0.92,1.04)* | *1.13 (0.99,1.30)* | **0.70 (0.60,0.83)** |
| Diabetes | 12.24 (11.33,13.21) | 13.94 (12.74,15.26) | 9.37 (8.46,10.37) | 18.33 (16.90,19.90) | 17.01 (14.11,20.50) | 9.81 (8.57,11.22) |
| Asthma | 2.10 (1.94,2.27) | 1.74 (1.61,1.87) | 1.69 (1.56,1.84) | 1.69 (1.52,1.89) | 1.83 (1.59,2.11) | 2.65 (2.08,3.39) |
| Gallbladder disease | 3.95 (3.70,4.22) | 2.44 (2.29,2.61) | 3.99 (3.69,4.31) | 3.31 (3.00,3.65) | 4.09 (3.24,5.16) | 2.54 (2.07,3.11) |
| Back pain | 1.44 (1.37,1.51) | 1.29 (1.23,1.35) | 1.27 (1.21,1.34) | 1.66 (1.55,1.79) | 1.31 (1.19,1.44) | 2.07 (1.77,2.42) |
| Osteoarthritis | 1.90 (1.82,1.98) | 1.51 (1.45,1.57) | 1.55 (1.48,1.62) | 2.67 (2.49,2.86) | 1.88 (1.69,2.10) | 3.58 (3.10,4.15) |

BIA bioimpedance analysis, BMI body mass index, CI confidence interval, lnCRP natural logarithm of C-reactive protein, SHR subdistribution hazard ratio, WHR waist:hip ratio

The models are adjusted for age, sex, lnCRP and a measure of body fat (linear and quadratic terms). Sex is interacted with the measures of body fat. Death is modelled as a competing risk. Significant negative associations are in blue font and bolded. Non-significant associations are in italics.

**Supplement Table S5. Associations of BMI or $fat_{BIA}$ and WHR with mortality and other clinical outcomes by sex, HR (95% CI) – 95th%ile vs 5th%ile**

| Outcome | Females | | Males | |
|---|---|---|---|---|
| | | *Model 1* | | |
| | *BMI* | *WHR* | *BMI* | *WHR* |
| Mortality | **0.75 (0.71,0.79)** | 1.82 (1.72,1.93) | **0.67 (0.64,0.71)** | 2.32 (2.21,2.45) |
| Cardiovascular | 2.36 (2.27,2.46) | 1.75 (1.69,1.82) | 1.87 (1.79,1.96) | 1.65 (1.58,1.72) |
| Cancer | 1.38 (1.29,1.46) | *0.98 (0.92,1.04)* | *0.94 (0.88,1.0005)* | 1.33 (1.25,1.42) |
| Diabetes | 5.82 (5.35,6.32) | 7.22 (6.56,7.94) | 5.26 (4.86,5.70) | 3.55 (3.25,3.89) |
| Asthma | 2.10 (1.91,2.30) | 1.60 (1.46,1.75) | *1.06 (0.95,1.19)* | 2.16 (1.92,2.42) |
| Gallbladder disease | 3.59 (3.34,3.87) | 1.66 (1.54,1.78) | 1.74 (1.58,1.91) | 2.29 (2.06,2.54) |
| Back pain | 1.59 (1.50,1.68) | 1.28 (1.22,1.35) | 1.39 (1.30,1.49) | 1.37 (1.27,1.46) |
| Osteoarthritis | 1.98 (1.88,2.09) | 1.26 (1.20,1.32) | 2.11 (1.97,2.26) | 1.24 (1.16,1.33) |
| | | *Model 2* | | |
| | *$Fat_{BIA}$* | *WHR* | *$Fat_{BIA}$* | *WHR* |
| Mortality | **0.73 (0.69,0.77)** | 1.81 (1.71,1.91) | **0.78 (0.74,0.82)** | 2.04 (1.93,2.15) |
| Cardiovascular | 1.89 (1.81,1.97) | 1.93 (1.86,2.01) | 1.50 (1.44,1.57) | 1.86 (1.77,1.94) |
| Cancer | 1.26 (1.18,1.34) | *1.02 (0.96,1.08)* | *0.94 (0.88,1.01)* | 1.32 (1.24,1.41) |
| Diabetes | 3.89 (3.50,4.32) | 9.11 (8.27,10.03) | 4.15 (3.77,4.57) | 4.29 (3.91,4.69) |
| Asthma | 1.79 (1.63,1.96) | 1.76 (1.61,1.92) | *1.09 (0.94,1.23)* | 2.15 (1.91,2.42) |
| Gallbladder disease | 3.98 (3.65,4.34) | 1.73 (1.62,1.86) | 2.18 (1.96,2.43) | 2.02 (1.81,2.25) |
| Back pain | 1.48 (1.40,1.56) | 1.33 (1.27,1.40) | 1.22 (1.14,1.31) | 1.49 (1.38,1.59) |
| Osteoarthritis | 1.59 (1.51,1.68) | 1.39 (1.32,1.46) | 1.41 (1.32,1.51) | 1.58 (1.47,1.70) |

BIA bioimpedance analysis, BMI body mass index, CI confidence interval, HR hazard ratio, lnCRP natural logarithm of c-reactive protein, WHR waist:hip ratio

The models are adjusted for age, sex, lnCRP, WHR and one other measure of body fat (linear and quadratic terms). The first model additionally adjusts for BMI. The second model additionally adjusts for $fat_{BIA}$. Sex is interacted with all measures of body fat. Significant negative associations are in blue font and bolded. Non-significant associations are in italics.
